# Multi-trait genetic analysis identifies novel pleiotropic loci for depression and schizophrenia in East Asians

**DOI:** 10.1101/2024.01.30.24301991

**Authors:** Yingchao Song, Linzehao Li, Yue Jiang, Bichen Peng, Hengxuan Jiang, Zhen Chao, Xiao Chang

**Author notes:** Correspondence to Xiao Chang, PhD, 6699 Qingdao Road, Jinan, China.

## Abstract

While genetic correlations, pleiotropic loci, and shared genetic mechanisms of psychiatric disorders have been extensively studied in European populations, the investigation of these factors in East Asian populations has been relatively limited. To identify novel pleiotropic risk loci for depression and schizophrenia (SCZ) in East Asians. We harnessed the most comprehensive dataset available for East Asians and quantified the genetic overlap between depression, SCZ, and their related traits via LD Score regression (LDSC) analyses. Besides the correlation between depression and SCZ, our analysis revealed significant genetic correlations between depression and obesity-related traits, such as weight, BMI, T2D, and HDL. In SCZ, significant correlations were detected with HDL, heart diseases and use of various medications. Conventional meta-analysis of depression and SCZ identified a novel locus at 1q25.2 in East Asians. Moreover, this locus was verified in the multi-trait analysis of GWAS (MTAG), which can improve the statistical power of single-trait GWAS by incorporating information from effect estimates across genetically correlated traits. Furthermore, multi-trait analysis of depression, SCZ and related traits identified ten novel pleiotropic loci for depression, and four for SCZ. Our findings demonstrate shared genetic underpinnings between depression and SCZ in East Asians, as well as their associated traits, providing novel candidate genes for the identification and prioritization of therapeutic targets specific to this population.

## Introduction

Psychiatric disorders comprise a range of severe illnesses that result in emotional, cognitive, or behavioral disturbances in individuals, making them a significant contributor to the global burden of disability [1]. Empirical studies indicate that psychiatric disorders result from the intricate interplay of genetic and environmental risk factors [2]. Heritability estimates, as determined by twin-based and genome-wide association studies (GWAS), emphasize the substantial role of genetic factors within the spectrum of psychiatric disorders [3–6]. Specifically, the heritability estimates for major depression are approximately 30%-40% [7], while for schizophrenia (SCZ), these estimates range from 65% to 80% [8–10].

Depression and SCZ both exhibit a substantial degree of polygenicity, with numerous risk loci have been identified through genome-wide association studies (GWAS) in individuals of European ancestry [6, 11]. In a recent study, the largest GWAS of depression, involving more than 1.3 million individuals (including 371,184 with depression), identified a total of 243 risk loci. This study also unveiled a critical role of neurodevelopmental pathways associated with prenatal GABAergic neurons, astrocytes and oligodendrocyte lineages [12]. For SCZ, a total of 287 significant loci have been identified in the largest GWAS encompassing 76,755 cases and 243,649 controls. These identified associations predominantly cluster within genes that are actively expressed in both excitatory and inhibitory neurons within the central nervous system [13]. However, the majority of previous genetic studies have been performed in European ancestry cohorts, leaving a conspicuous research gap in Asian populations. The largest GWAS of depression in East Asians (15,771 cases and 178,777 controls) only identified two genetic loci [14]. Similarly, the largest GWAS conducted on SCZ among individuals of East Asian origin (with 22,778 cases and 35,362 controls) identified 19 genetic loci [15], which is significantly fewer than those observed in European studies.

Recently, the emergence of multi-trait analysis of GWAS (MTAG) has brought forth a powerful approach to explore the genetic overlap and shared causality among complex traits [16]. By conducting meta-analysis of GWAS summary statistics for different genetically correlated traits, multi-trait analysis can enhance the statistical power and pinpoint pleiotropic genetic variants that influence interrelated traits. Moreover, it reveals the intricate genetic interplay existing between these phenotypes and unveils the common biological pathways that underpin their development. Such cross-trait analysis methods have been successfully applied to the study of psychiatric disorders. For example, a multi-trait meta-analysis across eight psychiatric disorders identified 109 pleiotropic loci linked to at least two disorders, revealing the common genetic structures shared among different psychiatric disorders [6]. Besides, a multi-trait meta-analysis revealed a substantial number of pleiotropic genetic loci between psychiatric disorder and gastrointestinal tract diseases, emphasizing the shared biological mechanism concerning immune response, synaptic structure and function, and potential gut microbiome in these conditions [17]. To the best of our knowledge, multi-trait analysis of GWAS has predominantly centered on European populations, and, no such analysis has been undertaken for psychiatric disorders within East Asian populations so far.

In this study, we conducted a comprehensive multi-trait analysis using GWAS data of depression and SCZ in cohorts of East Asian descent. We also investigated the deep-phenotype GWAS data encompassing a range of health aspects such as diseases, biomarkers and medication usage sourced from BioBank Japan (BBJ) [18]. Moreover, we performed analyses to quantify both overall and local genetic correlations, further identifying novel pleiotropic loci for depression and SCZ in East Asians.

## Methods

### GWAS data

The hitherto largest GWAS meta-analysis of depression among individuals of East Asia was performed among 194,548 individuals (15,771 cases and 178,777 controls), based on combined data from the China, Oxford, and Virginia Commonwealth University Experimental Research on Genetic Epidemiology (CONVERGE) consortium [19], China Kadoorie Biobank (CKB), and the Taiwan-Major Depressive Disorder (MDD) study, as well as studies conducted in the US and UK that included participants of East Asian ancestry [14]. And the largest GWAS of SCZ among individuals across East Asia were compiled 22,778 SCZ cases and 35,362 controls from 20 sample collections from East Asia [15].

GWAS summary statistics of 96 phenotypes from East Asians including diseases, biomarkers and medication usage were downloaded from GWAS Catalog [20], information of all included GWAS data from East Asians is summarized in Table S1.

### Meta-analysis between depression and SCZ

Inverse-variance weighted meta-analysis was performed on depression and SCZ using the basic meta-analysis function in PLINK (v1.90b7) and the fixed-effect meta-analysis p value and fixed-effect ORs were estimated. We prioritized significant SNPs that reached genome-wide significance (P < 5×10^−8^) in the meta-analysis and suggestive significance (P < 0.01) in the original single-trait GWAS.

### Global genetic correlation analysis

Genetic correlation r_g_ between depression and SCZ was estimated by LD (Linkage Disequilibrium) score regression (LDSC) using GWAS summary statistic [21]. The LDSC method is described by the following equation: 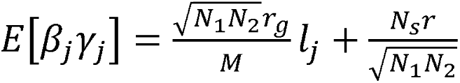, where *β_j_* and *γ_j_* denote the effect size of SNP *j* on the two tested traits, *N*_1_ and *N*_2_ are the sample sizes of two tested traits, *N_s_* is the number of overlapping samples between two tested traits, *r* is the phenotypic correlation in overlapping samples and *l_j_* is the LD score. Pre-computed linkage disequilibrium scores for HapMap3 SNPs calculated based on East-Asian-ancestry data from the 1000 Genomes Project were used in the analysis.

### Local genetic correlation analysis

Considering that genetic correlation estimated by LDSC aggregates information across all variants in the genome, we further estimated the pairwise local genetic correlation using ρ-HESS (heritability estimation from summary statistics) [22]. ρ-HESS are designed to quantify the local genetic correlation between pairs of traits at each of the 1703 prespecified LD independent segments with an average length of 1.6 Mb. A Bonferroni corrected p value less than 0.05/1703 was considered as statistically significant.

### Multi-trait GWAS analysis

In addition to the above traditional meta-analysis method, the Multi-Trait Analysis of GWAS (MTAG) framework, a generalized meta-analysis method that outputs trait-specific SNP associations [16], was furthered conducted. MTAG can increase the power to detect loci from correlated traits by analyzing GWAS summary statistics jointly. It also accounts for sample overlap and incomplete genetic correlation, when comparing with the conventional inverse-variance weighted meta-analysis. The first step of MTAG is to filter variants by removing non common SNPs, duplicated SNPs, or SNPs with strand ambiguity. MTAG then estimates the pairwise genetic correlation between traits using LDSC [23] and uses these estimates to calibrate the variance-covariance matrix of the random effect component. MTAG next performs a random-effect meta-analysis to generate the SNP-level summary statistics. We prioritized significant pleiotropic SNPs that reached genome-wide significance (*P* < 5×10^−8^) in the multi-trait analysis and suggestive significance (*P* < 0.01) in the original single-trait GWAS.

### Genetic correlation analysis between psychiatric disorders and deep phenotypes

To investigate the potential risk factors for psychiatric disorders in East Asian populations, we included 96 GWAS summary statistics of deep phenotypes from the BioBank Japan (BBJ), including diseases, biomarkers and medication usage. Genetic correlation r_g_ between depression/SCZ and each phenotype was estimated by LD (Linkage Disequilibrium) score regression (LDSC) using GWAS summary statistic [21].

### Multi-trait GWAS analysis of psychiatric disorders and risk factors

Multi-trait GWAS meta-analysis for psychiatric disorders and the corresponding genetically correlated deep phenotypes were performed by the MTAG framework. We prioritized significant pleiotropic SNPs that reached genome-wide significance (*P* < 5×10^−8^) in the multi-trait analysis and suggestive significance (P < 0.01) in the original single-trait GWAS.

## Results

### Meta-analysis between depression and schizophrenia

We conducted a comprehensive meta-analysis by combing summary statistics derived from the largest GWAS of depression and SCZ within East Asian populations. Our analysis yielded three pleiotropic loci surpassing the significance threshold of *P* < 5×10^−8^. Among them, 2p16.1 (rs13016665) and 18q23 (rs58736086) loci were previously reported in the study of SCZ among East Asian populations [15], while none of them was identified in the GWAS of depression (Table 1, Figure S1). The lead variant rs12031894 of the only novel locus is located in the intergenic region near gene *SOAT1*. It is worth mentioning that this locus reached genome-wide significance in the meta-analysis of SCZ data including both European and East Asian samples suggesting its potential relevance and impact in psychiatric disorders across diverse populations [15]. Additionally, the 1q25.2 locus (rs12031894) was identified as an expression quantitative trait locus (eQTL) for neighboring genes across multiple tissues (Table S2), particularly in brain tissues for genes *ABL2*, *FAM20B*, and *TDRD5* (Figure S2).

**Table 1.** Meta-analysis between depression and schizophrenia in East Asians using PLINK.

| SNP | CHR | BP | A1 | A2 | Depression |  | Schizophrenia |  | Meta |  | Genes | Annotation |
| --- | --- | --- | --- | --- | --- | --- | --- | --- | --- | --- | --- | --- |
|  |  |  |  |  | OR | *P*-value | OR | *P*-value | OR | *P*-value |  |  |
| rs12031894 | 1 | 179270314 | T | C | 0.948 | 0.001 | 0.928 | 1.24E-07 | 0.937 | 8.09E-10 | SOAT1 | new |
| rs13016665 | 2 | 57995348 | A | C | 1.058 | 4.11E-04 | 1.077 | 3.25E-06 | 1.067 | 7.03E-09 | VRK2 | known |
| rs58736086 | 18 | 77645575 | T | C | 1.066 | 0.003 | 1.097 | 2.30E-07 | 1.084 | 1.28E-08 | SLC66A2 | known |

### Genetic correlation between depression and schizophrenia

To further investigate the shared genetic mechanism between depression and SCZ in East Asian populations, we estimated the genetic correlation between the two psychiatric disorders using LDSC. In consistent with findings in European populations [5, 15, 24], we observed a significant positive genetic correlation between depression and SCZ in East Asian populations (r_g_ = 0.46, *P* = 1.48 × 10^-6^). In addition, heritability estimates on the observed scale using GWAS summary statistics of East Asians were 47.05% for SCZ which is comparable to the estimates calculated from Europeans. However, in the case of depression, heritability estimates among East Asians were notably lower at 0.78%, likely attributed to the relatively limited sample size of cases as compared to European studies.

### Multi-trait analysis between depression and schizophrenia

We next applied MTAG to detect the potential pleiotropic loci shared between depression and SCZ in East Asians. MTAG differs from conventional meta-analysis in its capacity to enhance the statistical power of GWAS by incorporating information from effect estimates across genetically correlated traits, and generate trait-specific associations of each single nucleotide polymorphism (SNP). In assessing the augmented detection power, we compared the average χ^2^ test statistic for depression or SCZ derived from the multi-trait GWAS with that originating from the initial GWAS conducted in East Asian populations. The mean χ^2^ statistics for the initial GWAS results are: χ^2^ _GWAS-DEP_ = 1.027 and χ^2^_GWAS-SCZ_ = 1.265 and the mean χ^2^ statistics for the MTAG-GWAS results are: χ^2^ _MTAG-DEP_ = 1.052 and χ^2^_MTAG-SCZ_ = 1.267. We observed varying degrees of increase in the effective sample size for both depression and SCZ. Specifically, the effective sample size for SCZ (MTAG-SCZ) increased slightly from 58,140 to 58,759, while the effective sample size for depression (MTAG-DEP) experienced a substantial estimated increase of 94.61%, expanding from 194,548 to 378,607. In alignment with the conventional meta-analysis conducted using PLINK, the newly identified signal at 1q25.2 was observed in both the results of the MTAG-DEP and MTAG-SCZ analyses (Table S3, Figure S1). We also detected two additional loci at 2q33.1 (rs17590956) and 18q23(rs28735056), which were previously reported in East Asians (Table S3, Figure S1).

### Genetic correlations between psychiatric disorders and deep phenotypes

To further elucidate the genetic correlations between psychiatric disorders and potential risk factors in East Asians, we analyzed GWAS data of 96 phenotypes sourced from BBJ, ranging from diseases, biomarkers and medication usage. Our analysis uncovered significant positive genetic correlations between depression and obesity-related traits. This encompassed variables like weight (r_g_ = 0.16, *P* = 0.009), body mass index (BMI, r_g_ = 0.24, *P* = 8.0 × 10^-4^), type 2 diabetes (T2D, r_g_ = 0.24, *P* = 0.003), and the use of medications associated with diabetes (r_g_ = 0.19, *P* = 0.022). Besides, serum alkaline phosphatase levels also exhibited a significant positive genetic correlation with depression. Conversely, our analysis unveiled significant negative genetic correlations between depression and two blood biomarkers: high-density lipoprotein (HDL) cholesterol and total bilirubin levels (Table S4, Figure 1A). Likewise, we detected significant negative correlations between SCZ and certain blood lipid markers, particularly HDL (r_g_ = -0.13, *P* = 0.006) and total cholesterol levels (TCL, r_g_ = -0.12, *P* = 0.018). In addition, significant negative genetic correlations were also detected between SCZ and chronic diseases including Chronic hepatitis C infection and Chronic sinusitis. In contrast, SCZ was significantly correlated with heart diseases such as angina pectoris. We also found significant genetic correlations between SCZ and the use of various medications, including those for peptic ulcer and gastro-esophageal reflux disease (GORD) (r_g_ = 0.34, *P* = 0.003), non-steroidal anti-inflammatory and antirheumatic products (r_g_ = 0.29, *P* = 0.012), among others (Table S4, Figure 1B).

**Figure 1.**
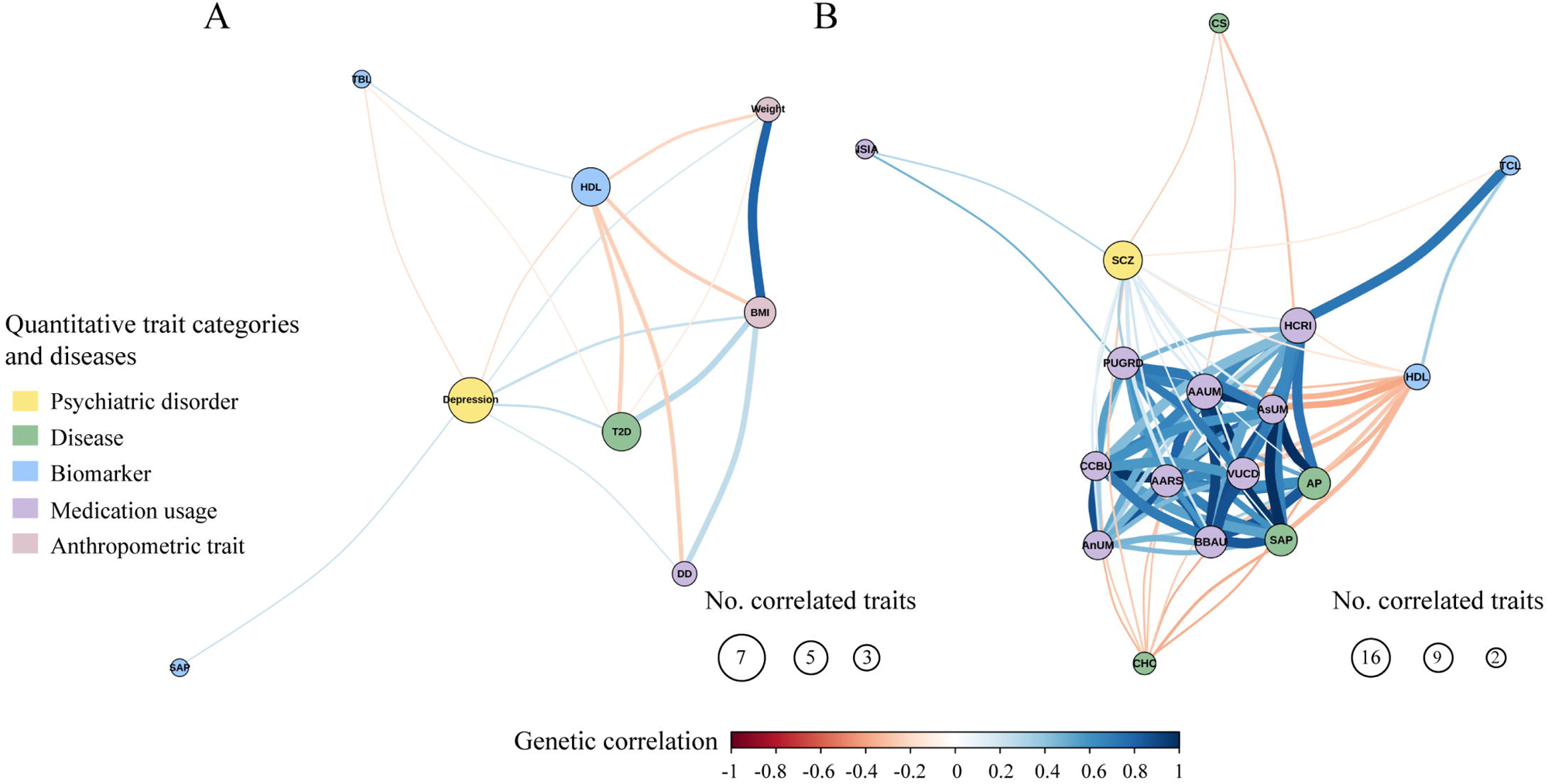
Genetic correlation network between psychiatric disorders and risk factors. (A) genetic correlation network of depression. (B) genetic correlation network of SCZ. Genetically correlated traits and diseases are clustered close together. Each circle represents a trait, and each edge represents a significant genetic correlation (PlJ<lJ0.05). Positive and negative genetic correlations are indicated by color according to the key. SCZ, SCZ; SAP (biomarker), serum alkaline phosphatase levels; TBL, total bilirubin levels; T2D, Type 2 diabetes; DD, Drugs used in diabetes use measurement; TCL, total cholesterol levels; AP, angina pectoris; CHC, chronic hepatitis C infection Chronic sinusitis; SAP (disease), stable angina pectoris; AARS, agents acting on the renin-angiotensin system use measurement; NSIA, Non-steroidal anti-inflammatory and antirheumatic product use measurement; AnUM, Antihypertensive use measurement; AAUM, Antithrombotic agent use measurement; BBAU, Beta blocking agent use measurement; CCBU, Calcium channel blocker use measurement; PUGRD, Peptic ulcer and gastro-oesophageal reflux disease (GORD) drug use measurement; HCRI, HMG CoA reductase inhibitor use measurement; Asum, Aspirin use measurement; VUCD, Vasodilators used in cardiac diseases use measurement.

### Local genetic correlations between psychiatric disorders and corresponding risk factors

We next conducted a genome-wide scan to identify specific genomic regions that contribute to the shared heritability of genetically correlated traits. Despite the absence of statistically significant regions following Bonferroni correction, we observed that 12q23-q24 (*P_local_*= 0.0019) and 16q21(*P_local_* = 0.0024) displayed suggestive correlations between depression and HDL (Figure S3A). Additionally, 12q23 (*P_local_* = 0.0012) exhibited a suggestive correlation between SCZ and HDL (Figure S3B).

### Multi-trait analysis of psychiatric disorders and corresponding risk factors

The insights garnered from our genetic correlation analysis served as a catalyst for a more in-depth exploration, aiming to pinpoint shared pleiotropic genetic loci that influence both psychiatric disorders and associated risk factors. Here, we conducted MTAG analyses for each genetically correlated trait pair, and prioritized SNPs that reached genome-wide significance (*P* < 5×10^−8^). We identified two significant signals in the analysis of depression and BMI, neither of which had been previously reported in GWAS studies of depression within East Asian populations. Among them, locus at 19q13.32 near *GIPR* was also detected in the analysis of depression and weight. The lead SNP rs4802269 serves as an eQTL for the nearby gene *EML2* in tissues from cerebellar hemispheres. Furthermore, it functions as an eQTL for the nearby genes *DMPK*, *DMWD*, and *GIPR* in adipose tissues (Table S2), implying the potential relevance of this locus in regulating factors associated with both depression and weight. Additionally, in our exploration of the connection between depression and type 2 diabetes (T2D), we uncovered three shared loci, none of which had been previously identified in studies of depression within East Asian populations. Among these, the variant rs10974438 at 9p24.2, located within an intronic region of *GLIS3*, has been linked to mania, hypomania, bipolar disorder, or manic-depression in an analysis of UK Biobank data (UKB Neale v2, Table S5). Furthermore, rs11074451 at 16p12.3 serves as a splicing quantitative trait locus (sQTL, *P* = 1 × 10^-9^) for the nearby gene *GP2* in the pancreas according to GTEx, suggesting its potential role in the interplay between depression and T2D. In addition to the above three loci, a novel locus at 18p11.22 was identified when analyzing depression and the measurement of diabetes medication usage. Besides, we identified two novel loci at 12q24.12 (rs11066015) and 16q12.2 (rs35626903) during the analysis of depression and HDL. The locus at 12q24.12 (rs11066015) is specific to East Asian populations and encompasses the pivotal ALDH2 gene, which has been implicated in various metabolic quantitative traits. The rs35626903 variant, located at 16q12.2 within the intron of the *AMFR* gene, has been previously related to psychological or psychiatric problem in the study of Europeans [25]. We also identified two novel pleiotropic signals between depression and total bilirubin levels (Table 2, Figure S4).

**Table 2.** Multi-trait meta-analysis between depression and risk factors in East Asians.

| Risk factors | SNP | CHR | BP | A1 | A2 | Trait1 |  | Trait2 |  | MTAG |  | Genes | Annotation |
| --- | --- | --- | --- | --- | --- | --- | --- | --- | --- | --- | --- | --- | --- |
|  |  |  |  |  |  | beta | P-value | beta | P-value | beta | P-value |  |  |
| BMI | rs13029479 | 2 | 655222 | A | G | 0.101 | 0.007 | -0.052 | 5.40E-17 | 0.035 | 1.31E-09 | TMEM18 | new |
|  | rs4802269 | 19 | 46167469 | G | A | -0.07 | 4.94E-04 | 0.042 | 4.60E-16 | -0.021 | 5.01E-11 | GIPR | new |
| Weight | rs4802269 | 19 | 46167469 | G | A | -0.07 | 4.94E-04 | 0.037 | 4.30E-17 | -0.021 | 1.25E-08 | GIPR | new |
| HDL | rs11066015 | 12 | 112168009 | A | G | -0.073 | 0.001 | -0.122 | 1.60E-109 | -0.033 | 8.23E-21 | ALDH2 | new |
|  | rs35626903 | 16 | 56431613 | G | A | -0.071 | 6.24E-06 | -0.028 | 2.50E-09 | -0.018 | 2.52E-09 | AMFR | new |
| TBL | rs12993249 | 2 | 234479377 | A | G | -0.055 | 1.40E-03 | -0.123 | 1.60E-208 | -0.044 | 2.55E-46 | USP40 | new |
|  | rs10770745 | 12 | 20986617 | T | G | -0.067 | 8.73E-04 | -0.031 | 2.60E-13 | -0.018 | 9.50E-09 | SLCO1B3<br>-<br>SLCO1B7 | new |
|  | rs11066015 | 12 | 112168009 | A | G | -0.073 | 1.47E-03 | -0.035 | 1.30E-15 | -0.02 | 4.66E-09 | ALDH2 | new |
| Type 2 diabetes | rs10013800 | 4 | 71731236 | T | C | -0.092 | 4.21E-05 | 0.081 | 2.21E-10 | -0.023 | 1.19E-11 | GRSF1 | new |
|  | rs10974438 | 9 | 4291928 | A | C | 0.045 | 0.005 | 0.078 | 2.35E-16 | 0.017 | 4.95E-12 | GLIS3 | new |
|  | rs11074451 | 16 | 20336257 | A | G | 0.1 | 2.27E-04 | 0.103 | 9.12E-11 | 0.026 | 5.61E-11 | GP2 | new |
| Drugs used in diabetes use measurement | rs6850072 | 4 | 71725868 | A | G | 0.074 | 2.37E-04 | -0.086 | 1.14E-12 | 0.022 | 1.25E-11 | GRSF1 | new |
|  | rs10974438 | 9 | 4291928 | A | C | 0.045 | 0.005 | 0.072 | 1.91E-12 | 0.016 | 1.64E-09 | GLIS3 | new |
|  | rs11074451 | 16 | 20336257 | A | G | 0.1 | 2.27E-04 | 0.114 | 2.00E-11 | 0.028 | 5.46E-11 | GP2 | new |
|  | rs662872 | 18 | 8881068 | T | C | -0.084 | 1.80E-06 | 0.038 | 7.27E-04 | -0.016 | 2.02E-08 | MTCL1 | new |

Trait1: depression
Trait2: risk factors

We next conducted a MTAG analysis to uncover pleiotropic variants impacting both SCZ and corresponding risk factors. We identified a total of seven significant loci shared between SCZ and HDL including six located in loci reported in SCZ GWAS in East Asian populations. The novel locus at 5q31.3 was also identified in the analysis between SCZ and TCL, as well as the analysis between SCZ and stable angina pectoris (Table 3, Figure S5). Notably, this locus is sited close to the protocadherin gamma gene cluster, which may play a critical role in the establishment and function of specific cell-cell connections in the brain. Particularly, the lead SNP rs11956411 is an eQTL of *PCDHGA3* in brain cortex (Table S2). Additionally, we detected four significant loci shared between SCZ and chronic diseases (Chronic hepatitis C infection and Chronic sinusitis), but all of them were reported in GWAS of SCZ conducted on East Asians (Table S6, Figure S7-S8). In the analysis of SCZ and the use of medications, we uncovered three novel loci at 1q25.2 (rs61824368), 4q13.1 (rs62297721), and 8p21.2 (rs117325001) (Table 3, Figure S5). The 1q25.2 and 8p21.2 loci had been reported in the meta-analysis of SCZ, including both European and East Asian populations [15].

**Table 3.** Novel loci in the multi-trait meta-analysis between schizophrenia and risk factors in East Asians.

| Risk factors | SNP | CHR | BP | A1 | A2 | Trait1 |  | Trait2 |  | MTAG |  | Genes | Annotation |
| --- | --- | --- | --- | --- | --- | --- | --- | --- | --- | --- | --- | --- | --- |
|  |  |  |  |  |  | beta | P-value | beta | P-value | beta | P-value |  |  |
| HDL | rs11956411 | 5 | 140853856 | C | T | -0.097 | 1.08E-07 | -0.02 | 1.40E-03 | -0.035 | 2.77E-08 | PCDHGC3 | new |
| TCL | rs11956411 | 5 | 140853856 | T | C | 0.097 | 1.08E-07 | 0.005 | 1.00E-03 | 0.036 | 1.54E-08 | PCDHGC3 | new |
| Stable angina pectoris | rs2237079 | 5 | 140877905 | A | G | -0.085 | 1.24E-07 | 0.013 | 1.94E-03 | -0.033 | 4.65E-08 | PCDHGC3 | new |
| Antihypertensive use measurement | rs62297721 | 4 | 66504803 | A | G | 0.074 | 6.44E-08 | 0.019 | 4.81E-03 | 0.033 | 1.78E-08 | EPHA5 | new |
| Antithrombotic agent use measurement | rs117325001 | 8 | 26242272 | T | G | 0.074 | 6.62E-08 | 0.008 | 8.98E-03 | 0.033 | 2.49E-08 | BNIP3L | new |
| aspirin use measurement | rs117325001 | 8 | 26242272 | T | G | 0.074 | 6.62E-08 | 0.008 | 9.52E-04 | 0.033 | 2.44E-08 | BNIP3L | new |
| Vasodilators used in cardiac diseases use measurement | rs61824368 | 1 | 179298809 | T | C | -0.075 | 1.19E-07 | 0.013 | 2.51E-03 | -0.033 | 3.26E-08 | SOAT1 | new |

Trait1: depression
Trait2: risk factors

## Discussion

To the best of our current understanding, this is the first genome-wide multi-trait analysis investigating genetic correlations, pleiotropic loci and shared genetic mechanisms of depression and SCZ within East Asian populations. Leveraging the known genetic correlation between these two psychiatric disorders, we pinpointed a novel locus via the inverse-variance weighted meta-analysis, and further validated this finding using MTAG. To explore the potential risk factors for psychiatric disorders among East Asian populations, we conducted an in-depth examination of 96 distinct phenotypes from BBJ. This extensive exploration yielded the identification of several previously unreported risk loci for SCZ and depression, shedding new light on the distinctive genetic underpinnings of these psychiatric disorders within the East Asian populations.

In previous research efforts conducted by the Psychiatric Genomics Consortium (PGC), conventional meta-analysis techniques were utilized to detect pleiotropic loci associated with psychiatric disorders, with a primary focus on European populations [6, 26]. However, recent progress in GWAS focused on SCZ and depression has led to a substantial increase in data concerning East Asian populations, which motivated us to initiate research endeavors focused exclusively on East Asian populations. Our meta-analysis of depression and SCZ identified a novel locus at 1q25.2 shared between depression and SCZ in East Asians. The lead single nucleotide polymorphism (SNP), rs12031894, has been identified as an eQTL influencing the activity of multiple nearby genes across various tissues. Notably, in a majority of brain tissues, rs12031894 functions as an eQTL for *ABL2*. *ABL2* encodes a member of the Abelson family of nonreceptor tyrosine protein kinases, which play pivotal roles in processes such as neurulation and neurite outgrowth. These kinases act as intermediaries, transmitting signals from axon guidance cues and growth factor receptors to facilitate cytoskeletal rearrangements. In mature neurons, Abl kinases are situated in both pre- and postsynaptic compartments, contributing to the regulation of synaptic stability and plasticity. Given that this locus exhibits an association with SCZ in European populations as well (Figure 2A), it suggests that the candidate gene *ABL2* could exerts a widespread influence on SCZ, affecting both East Asians and Europeans. In contrast, the evidence from regional plot (Figure 2B) suggests that this locus might be exclusively linked to depression in East Asians but not in Europeans, implying the existence of a complex regulatory mechanism influencing depression across different populations. We then utilized MTAG to identify shared pleiotropic loci between depression and SCZ in East Asians. MTAG enhances GWAS statistical power by integrating effect estimates from genetically related traits to derive trait-specific SNP associations, distinguishing it from conventional meta-analysis. As expected, the aforementioned 1q25.2 locus appeared in both MTAG-DEP and MTAG-SCZ results, demonstrating the reliability of this novel method.

**Figure 2.**
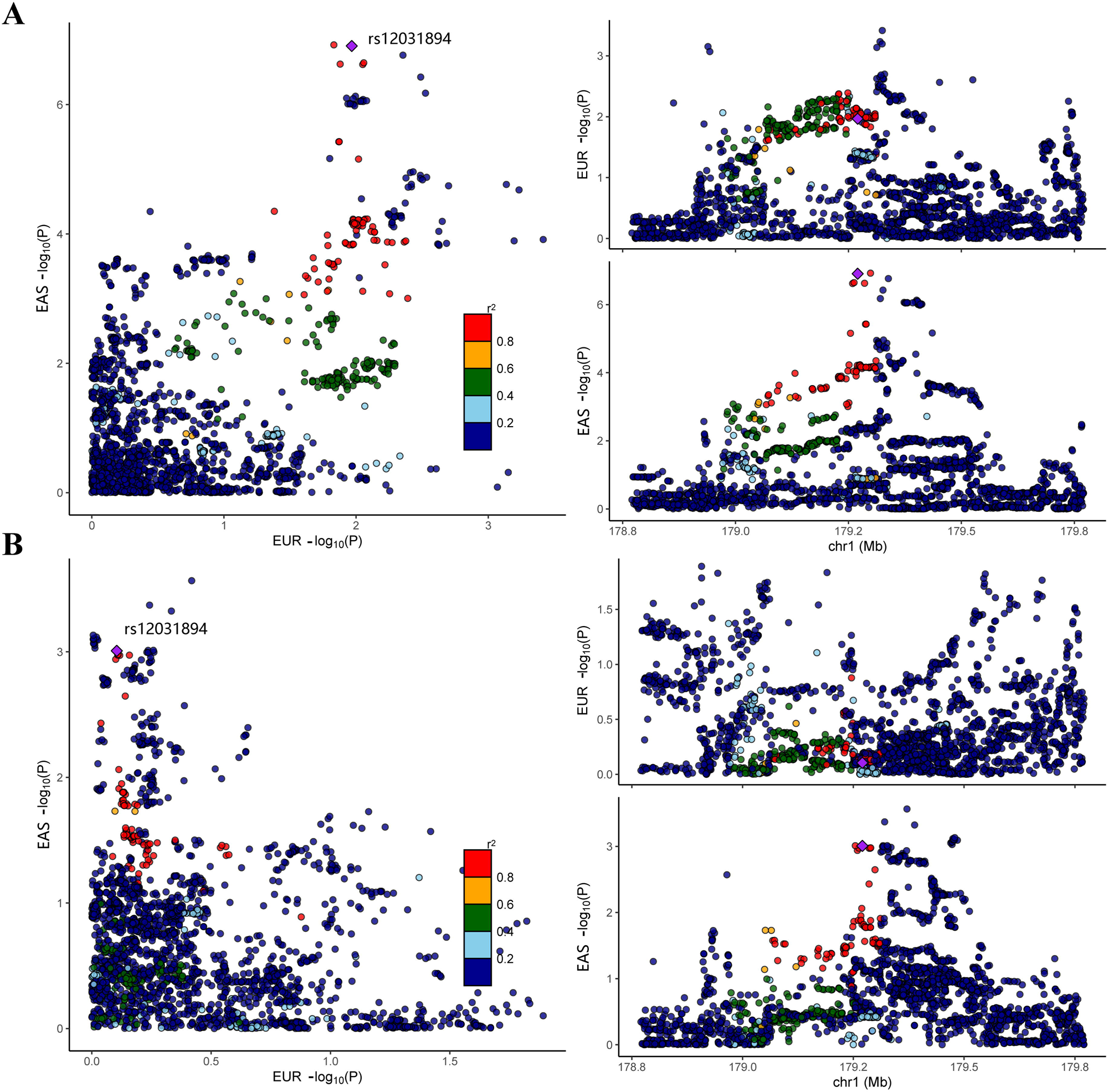
Co-localization for psychiatric disorders in East Asians and Europeans at 1q25.2. Left panel showed the merged association plot for psychiatric disorder in East Asians and Europeans, and right panel showed the regional plots for East Asians and Europeans respectively. Panel A shows the regional plots for SCZ and points are colored by LD with respect to rs12031894, which is labeled with a purple diamond. Panel B shows the regional plots for depression and points are colored by LD with respect to rs12031894, which is labeled with a purple diamond.

In addition to correlations among psychiatric disorders [27], LDSC analysis has consistently reported genetic correlations between psychiatric disorders and other diseases or risk factors in European populations [28, 29]. Therefore, we extended our investigations to East Asian populations by analyzing GWAS data encompassing 96 phenotypes from the BBJ dataset. Our analysis revealed positive genetic correlations between depression and obesity-related traits, such as weight, BMI, T2D, and diabetes medication usage. This aligns with findings from various epidemiological studies [30–33]. Intriguingly, serum alkaline phosphatase levels also displayed a positive genetic correlation with depression. This is consistent with the observations of Li et al., who reported higher alkaline phosphatase levels in individuals with depression compared to a control group in their analysis of National Health and Nutrition Examination Survey (NHANES) data [34]. Furthermore, recent research on U.S. adults has indicated a significant association between higher alkaline phosphatase levels and an increased risk of depression [35]. Conversely, we identified negative genetic correlations between depression and two blood biomarkers: HDL and bilirubin. Low levels of HDL have been extensively studied in connection with depression in multiple research studies [36–38]. Notably, Liang et al. reported that low HDL levels may be linked to more severe depressive symptoms in East Asian populations [39]. Additionally, lower HDL levels have been associated with SCZ and psychosis in various studies [40–42]. Consistent with this, we observed negative genetic correlations between HDL and SCZ. In contrast, positive genetic correlations were noted with heart diseases such as angina pectoris, which is in line with the observation that cardiovascular diseases are prevalent among individuals with SCZ [43–45]. These genetic correlations provide valuable insights into the intricate interplay among depression, SCZ, metabolic factors, and various health conditions in East Asian populations, guiding further research and facilitating the identification of shared pleiotropic loci influencing both psychiatric disorders and related risk factors.

We next performed MTAG analysis on depression, SCZ, and their associated traits in East Asian populations. We uncovered ten novel pleiotropic loci for depression through MTAG analysis. These loci demonstrated connections with traits such as BMI, weight, HDL, TBL, T2D, and diabetes medications. Seven of these loci exhibited evidence of association with psychiatric traits in Europeans, including four acting as eQTLs, implying their potential regulatory role in gene expression. For instance, the lead SNP rs4802269 of the 19q13.32 locus served as an eQTL for the nearby gene *EML2* in cerebellar hemispheres and for genes *DMPK*, *DMWD*, and *GIPR* in adipose tissues, hinting at its role in depression and weight regulation. Additionally, 18p11.22 was identified in the analysis of depression and diabetes medication usage. This locus is near the *MTCL1* gene, which plays a vital role in maintaining Purkinje neuron axon initial segments [46]. Furthermore, among the remaining genetic loci, the 12q24.12 locus is specifically presented in East Asian populations. Notably, the lead variant rs11066015 within this region exhibits a high level of LD with a missense variant of gene *ALDH2* (rs671, R^2^ = 0.95). Our analysis reveals that the MTAG P-value for rs671 (*P* = 4.46×10^-20^) is slightly greater than that of rs11066015 (*P* = 8.23×10^-21^), suggesting that the 12q24.12 locus is likely influenced by rs671 in East Asians. This is of particular interest because the rs671 variant leads to a loss of ALDH2 enzymatic activity, resulting in adverse reactions to acetaldehyde, which effectively reduce alcohol consumption [47].

In the investigation of SCZ and its related traits, we uncovered four new genetic loci, one of which was the previously mentioned 1q25.2 locus, shared between depression and SCZ. Notably, the 5q31.3 locus emerged consistently in the analysis of SCZ and its associated traits, which included HDL, TCL, and Stable angina pectoris. This locus is situated in close proximity to the protocadherin gamma gene cluster, which plays a potentially vital role in establishing and maintaining specific cell-cell connections in the brain. Particularly, the lead SNP rs11956411 acts as an eQTL for *PCDHGA3* in the brain cortex. Moreover, the 4q13.1 locus is located around the *EPHA5* gene, known for its crucial involvement in brain development and synaptic plasticity. This locus may also be linked to brain disorders such as bipolar [48] and Dravet syndrome [49]. Similar to the previously mentioned 1q25.2 locus, the 8p21.2 locus centered around the *BNIP3L* gene surpassed genome-wide significance in the meta-analysis of SCZ GWAS from both European and East Asian populations. Further investigation of both common and rare genetic variants indicated that *BNIP3L* might serve as a susceptibility gene [50]. However, it’s important to note that eQTL data from GTEx suggests that genotypes of the lead SNP rs117325001 are significantly associated with the expression level of *SDAD1P1* in multiple brain tissues, implying a potential role of *SDAD1P1* in SCZ.

This study has limitations. Firstly, due to the constraints of available data within the East Asian population, our analysis was limited to depression and SCZ. As more comprehensive datasets become accessible, we will have the opportunity to explore and analyze a broader spectrum of psychiatric disorders in East Asians, including but not limited to ADHD, Autism, and Anorexia, which will contribute to a more comprehensive understanding of genetic correlations across various mental health conditions. Secondly, it’s important to acknowledge that the eQTL data utilized in this study primarily stem from non-East Asian populations, predominantly Europeans. As a result, the potential cis-eQTL effects observed should ideally be subjected to further validation within East Asian populations once such data becomes available. This verification process will enhance the reliability and generalizability of our findings, ensuring that they are representative of the genetic landscape in East Asian individuals.

In conclusion, our study advances our understanding of the genetic mechanisms of depression and SCZ within East Asian populations by introducing 14 previously undiscovered pleiotropic risk loci to the existing pool of genetic contributors to psychiatric disorders. It is evident from this study that additional genetic loci associated with psychiatric disorders in East Asian populations can be identified through standard GWAS or MTAG analysis with larger sample sizes. This study warrants further investigation to explore the genetic intricacies of mental health conditions within this specific population.

## Data Availability

All data produced in the present study are available upon reasonable request to the authors

## Acknowledgements

This work is supported by the Special Funds of Taishan Scholar Project, China (tsqn202211224), All authors report no biomedical financial interests or potential conflicts of interest.

