## Supplemental Tables and Figures for "Multi-trait genetic analysis identifies novel pleiotropic loci for depression and schizophrenia in East Asians"

**Song *et al.***

**Supplementary Online Contents**

**Table S1.** Details on the characteristics of GWAS datasets in this study.

**Table S2.** Single-Tissue eQTLs for novel loci.

**Table S3.** Multi-trait analysis between depression and schizophrenia in East Asians using MTAG.

**Table S4.** Significant genetic correlations between psychiatric disorders and deep phenotypes.

**Table S5.** Diseases and traits associated with novel identified loci in MTAG.

**Table S6.** Known loci in multi-trait meta-analysis between schizophrenia and risk factors in East Asians.

**Figure S1.** Manhattan plot of the GWAS meta-analysis between depression and SCZ.

**Figure S2.** Multi-tissue eQTL plot for rs12031894.

**Figure S3.** Local genetic correlation between psychiatric disorders and HDL cholesterol in East Asians.

**Figure S4.** Manhattan plot of the GWAS multi-trait meta-analysis between depression and risk factors.

**Figure S5.** Manhattan plot of the GWAS multi-trait meta-analysis between SCZ and risk factors.

**Figure S6.** Manhattan plot of multi-trait meta-analysis between schizophrenia and angina pectoris in East Asians.

**Figure S7.** Manhattan plot of multi-trait meta-analysis between schizophrenia and chronic hepatitis C infection in East Asians.

**Figure S8.** Manhattan plot of multi-trait meta-analysis between schizophrenia and chronic sinusitis in East Asians.

**Figure S9.** Manhattan plot of multi-trait meta-analysis between schizophrenia and agents acting on the renin-angiotensin system use measurement in East Asians.

**Figure S10.** Manhattan plot of multi-trait meta-analysis between schizophrenia and non-steroidal anti-inflammatory and antirheumatic product use measurement in East Asians.

**Figure S11.** Manhattan plot of multi-trait meta-analysis between schizophrenia and beta blocking agent use measurement in East Asians

**Figure S12.** Manhattan plot of multi-trait meta-analysis between schizophrenia and calcium channel blocker use measurement in East Asians.

**Figure S13.** Manhattan plot of multi-trait meta-analysis between schizophrenia and peptic ulcer and gastro-oesophageal reflux disease (GORD) drug use measurement in East Asians.

**Figure S14.** Manhattan plot of multi-trait meta-analysis between schizophrenia and HMG CoA reductase inhibitor use measurement in East Asians.

**Table S1.** Details on the characteristics of GWAS datasets in this study.

| **Phenotype** | **Data source** | **Total sample size** | **Ncase** | **Ncontrol** | **GWAS Catalog accession ID** |
| --- | --- | --- | --- | --- | --- |
| Depression | PMID: 34586374 | 194548 | 15771 | 178777 |  |
| Schizophrenia | PMID: 31740837 | 58140 | 22778 | 35362 |  |
| Body mass index | PMID: 34594039 | 163,835 |  |  | GCST90018727 |
| HDL cholesterol | PMID: 34594039 | 74,970 |  |  | GCST90018736 |
| Serum alkaline phosphatase levels | PMID: 34594039 | 118886 |  |  | GCST90018722 |
| Total bilirubin levels | PMID: 34594039 | 124,341 |  |  | GCST90018753 |
| Total cholesterol levels | PMID: 34594039 | 135808 |  |  | GCST90018754 |
| Weight | PMID: 34594039 | 165419 |  |  | GCST90018729 |
| Agents acting on the renin-angiotensin system use measurement | PMID: 34594039 | 178726 | 45,820 | 132,906 | GCST90018768 |
| Non-steroidal anti-inflammatory and antirheumatic product use measurement | PMID: 34594039 | 178726 | 29694 | 149032 | GCST90018772 |
| Antihypertensive use measurement | PMID: 34594039 | 178726 | 5688 | 173038 | GCST90018764 |
| Antithrombotic agent use measurement | PMID: 34594039 | 178726 | 54220 | 124506 | GCST90018762 |
| Beta blocking agent use measurement | PMID: 34594039 | 178726 | 20367 | 158359 | GCST90018766 |
| Calcium channel blocker use measurement | PMID: 34594039 | 178726 | 49327 | 129399 | GCST90018767 |
| Peptic ulcer and gastro-oesophageal reflux disease (GORD) drug use measurement | PMID: 34594039 | 178726 | 53363 | 125363 | GCST90018760 |
| Drugs used in diabetes use measurement | PMID: 34594039 | 178726 | 30515 | 148211 | GCST90018761 |
| HMG CoA reductase inhibitor use measurement | PMID: 34594039 | 178726 | 33295 | 145431 | GCST90018769 |
| aspirin use measurement | PMID: 34594039 | 178726 | 41461 | 137265 | GCST90018775 |
| Vasodilators used in cardiac diseases use measurement | PMID: 34594039 | 178726 | 17050 | 161676 | GCST90018763 |
| Angina pectoris | PMID: 34594039 | 159165 | 14007 | 145158 | GCST90018573 |
| Chronic hepatitis C infection | PMID: 34594039 | 176698 | 7110 | 169588 | GCST90018585 |
| Chronic sinusitis | PMID: 34594039 | 156843 | 4617 | 152226 | GCST90018603 |
| Stable angina pectoris | PMID: 34594039 | 165047 | 18833 | 146214 | GCST90018695 |
| Type 2 diabetes | PMID: 34594039 | 177415 | 45383 | 132032 | GCST90018706 |
| Alanine aminotransferase levels | PMID: 34594039 | 150545 |  |  | GCST90018723 |
| Aspartate aminotransferase levels | PMID: 34594039 | 150068 |  |  | GCST90018724 |
| Basophil count | PMID: 34594039 | 91908 |  |  | GCST90018726 |
| Blood urea nitrogen levels | PMID: 34594039 | 148767 |  |  | GCST90018728 |
| C-reactive protein | PMID: 34594039 | 83025 |  |  | GCST90018730 |
| Calcium levels | PMID: 34594039 | 83980 |  |  | GCST90018731 |
| Diastolic blood pressure | PMID: 34594039 | 145515 |  |  | GCST90018732 |
| Eosinophil counts | PMID: 34594039 | 93063 |  |  | GCST90018733 |
| Gamma glutamyl transpeptidase | PMID: 34594039 | 133471 |  |  | GCST90018734 |
| Glucose levels | PMID: 34594039 | 133336 |  |  | GCST90018735 |
| Height | PMID: 34594039 | 165056 |  |  | GCST90018739 |
| Hematocrit | PMID: 34594039 | 153015 |  |  | GCST90018740 |
| Hemoglobin | PMID: 34594039 | 152447 |  |  | GCST90018737 |
| Hemoglobin A1c levels | PMID: 34594039 | 71221 |  |  | GCST90018738 |
| LDL cholesterol | PMID: 34594039 | 72866 |  |  | GCST90018741 |
| Mean arterial pressure | PMID: 34594039 | 145502 |  |  | GCST90018743 |
| Mean corpuscular hemoglobin | PMID: 34594039 | 128028 |  |  | GCST90018744 |
| Mean corpuscular hemoglobin concentration | PMID: 34594039 | 135482 |  |  | GCST90018745 |
| Mean corpuscular volume | PMID: 34594039 | 129832 |  |  | GCST90018746 |
| Monocyte count | PMID: 34594039 | 95119 |  |  | GCST90018747 |
| Neutrophil count | PMID: 34594039 | 82810 |  |  | GCST90018748 |
| Platelet count | PMID: 34594039 | 148623 |  |  | GCST90018749 |
| Pulse pressure | PMID: 34594039 | 145445 |  |  | GCST90018750 |
| Red blood cell count | PMID: 34594039 | 153512 |  |  | GCST90018751 |
| Serum albumin levels | PMID: 34594039 | 120539 |  |  | GCST90018725 |
| Serum creatinine levels | PMID: 34594039 | 150266 |  |  | GCST90018759 |
| Serum total protein level | PMID: 34594039 | 133321 |  |  | GCST90018756 |
| Serum uric acid levels | PMID: 34594039 | 129405 |  |  | GCST90018757 |
| Systolic blood pressure | PMID: 34594039 | 145505 |  |  | GCST90018752 |
| Triglycerides | PMID: 34594039 | 111667 |  |  | GCST90018755 |
| White blood cell count | PMID: 34594039 | 154355 |  |  | GCST90018758 |
| Lymphocyte count | PMID: 34594039 | 95717 |  |  | GCST90018742 |
| Inhalant adrenergic use measurement | PMID: 34594039 | 178726 | 5880 | 172846 | GCST90018999 |
| Anilide use measurement | PMID: 34594039 | 178726 | 4421 | 174305 | GCST90018776 |
| Antidepressant use measurement | PMID: 34594039 | 178726 | 3288 | 175438 | GCST90018778 |
| Antiglaucoma preparations and miotics use measurement | PMID: 34594039 | 178726 | 5911 | 172815 | GCST90018782 |
| Antihistamine use measurement | PMID: 34594039 | 178726 | 10163 | 168563 | GCST90018781 |
| Diuretic use measurement | PMID: 34594039 | 178726 | 22356 | 156370 | GCST90018765 |
| Drugs affecting bone structure and mineralization use measurement | PMID: 34594039 | 178726 | 6143 | 172583 | GCST90018773 |
| Glucocorticoid use measurement | PMID: 34594039 | 178726 | 13102 | 165624 | GCST90018780 |
| Immunosuppressant use measurement | PMID: 34594039 | 178726 | 3483 | 175243 | GCST90018771 |
| Opioid use measurement | PMID: 34594039 | 178726 | 3566 | 175160 | GCST90018774 |
| Thyroid preparation use measurement | PMID: 34594039 | 178726 | 3103 | 175623 | GCST90018770 |
| Allergic conjunctivitis | PMID: 34594039 | 178619 | 3900 | 174719 | GCST90018571 |
| Allergic rhinitis | PMID: 34594039 | 161563 | 7897 | 153666 | GCST90018572 |
| Asthma | PMID: 34594039 | 175948 | 13015 | 162933 | GCST90018575 |
| Atopic dermatitis | PMID: 34594039 | 168103 | 4296 | 163807 | GCST90018564 |
| Atrial fibrillation/atrial flutter | PMID: 34594039 | 159690 | 4150 | 155540 | GCST90018576 |
| Breast cancer | PMID: 34594039 | 79550 | 6325 | 73225 | GCST90018579 |
| Cataracts | PMID: 34594039 | 178726 | 38194 | 140532 | GCST90018594 |
| Cerebral aneurysm | PMID: 34594039 | 155154 | 3132 | 152022 | GCST90018595 |
| Cholelithiasis | PMID: 34594039 | 177558 | 9305 | 168253 | GCST90018599 |
| Chronic heart failure | PMID: 34594039 | 178726 | 10540 | 168186 | GCST90018586 |
| Chronic obstructive pulmonary disease | PMID: 34594039 | 166670 | 4017 | 162653 | GCST90018587 |
| Colon polyp | PMID: 34594039 | 170820 | 4768 | 166052 | GCST90018607 |
| Colorectal cancer | PMID: 34594039 | 167691 | 8305 | 159386 | GCST90018588 |
| Food allergy | PMID: 34594039 | 169716 | 3777 | 165939 | GCST90018625 |
| Gastric cancer | PMID: 34594039 | 167122 | 7921 | 159201 | GCST90018629 |
| Gastric ulcer | PMID: 34594039 | 173877 | 12650 | 161227 | GCST90018631 |
| Glaucoma | PMID: 34594039 | 177351 | 8448 | 168903 | GCST90018632 |
| Hearing loss, difficulty in hearing | PMID: 34594039 | 178726 | 3400 | 175326 | GCST90018637 |
| Iron deficiency anemia | PMID: 34594039 | 178675 | 4618 | 174057 | GCST90018652 |
| Ischemic stroke | PMID: 34594039 | 174686 | 22664 | 152022 | GCST90018644 |
| Lung cancer | PMID: 34594039 | 178726 | 4444 | 174282 | GCST90018655 |
| Myocardial infarction | PMID: 34594039 | 161206 | 14992 | 146214 | GCST90018657 |
| Osteoporosis | PMID: 34594039 | 178726 | 9794 | 168932 | GCST90018667 |
| Periodontal disease | PMID: 34594039 | 178726 | 9560 | 169166 | GCST90018677 |
| Peripheral artery disease | PMID: 34594039 | 177713 | 4112 | 173601 | GCST90018670 |
| Pneumonia | PMID: 34594039 | 178726 | 7423 | 171303 | GCST90018681 |
| Pollinosis | PMID: 34594039 | 172259 | 18593 | 153666 | GCST90018683 |
| Prostate cancer | PMID: 34594039 | 90332 | 5672 | 84660 | GCST90018685 |
| Rheumatoid arthritis | PMID: 34594039 | 178616 | 5348 | 173268 | GCST90018690 |
| Urolithiasis | PMID: 34594039 | 178726 | 11699 | 167027 | GCST90018715 |
| Uterine fibroids | PMID: 34594039 | 80208 | 14475 | 65733 | GCST90018714 |

**Table S2. Single-Tissue eQTLs for novel loci.**

| **Trait** | **Locus** | **SNP** | **CHR** | **BP** | **A1** | **A2** | **Gene Symbol** | **P-Value** | **NES** | **Tissue** |
| --- | --- | --- | --- | --- | --- | --- | --- | --- | --- | --- |
| Depression  &  Schizophrenia | 1q25.2 | rs12031894 | 1 | 179270314 | T | C | ABL2 | 5.50E-05 | 0.33 | [Brain - Anterior cingulate cortex (BA24)](javascript:gtex.goTissuePage('Artery_Tibial')) |
|  |  |  |  |  |  |  |  | 8.30E-07 | 0.31 | [Brain - Cerebellar Hemisphere](javascript:gtex.goTissuePage('Artery_Tibial')) |
|  |  |  |  |  |  |  |  | 2.50E-05 | 0.27 | [Brain - Cerebellum](javascript:gtex.goTissuePage('Nerve_Tibial')) |
|  |  |  |  |  |  |  |  | 1.10E-05 | 0.32 | [Brain - Cortex](javascript:gtex.goTissuePage('Skin_Sun_Exposed_Lower_leg')) |
|  |  |  |  |  |  |  |  | 7.10E-07 | 0.34 | [Brain - Frontal Cortex (BA9)](javascript:gtex.goTissuePage('Adipose_Subcutaneous')) |
|  |  |  |  |  |  |  |  | 1.40E-05 | 0.28 | [Brain - Hippocampus](javascript:gtex.goTissuePage('Thyroid')) |
|  |  |  |  |  |  |  |  | 1.60E-05 | 0.18 | [Brain - Nucleus accumbens (basal ganglia)](javascript:gtex.goTissuePage('Artery_Aorta')) |
|  |  |  |  |  |  |  |  | 3.10E-05 | 0.23 | [Brain - Putamen (basal ganglia)](javascript:gtex.goTissuePage('Cells_Cultured_fibroblasts')) |
|  |  |  |  |  |  |  |  | 3.00E-06 | 0.12 | [Cells - Cultured fibroblasts](javascript:gtex.goTissuePage('Esophagus_Mucosa')) |
|  |  |  |  |  |  |  | AXDND1 | 1.90E-06 | -0.2 | [Colon - Transverse](javascript:gtex.goTissuePage('Heart_Atrial_Appendage')) |
|  |  |  |  |  |  |  | FAM20B | 9.30E-07 | -0.11 | [Adipose - Subcutaneous](javascript:gtex.goTissuePage('Artery_Tibial')) |
|  |  |  |  |  |  |  |  | 4.40E-05 | -0.13 | [Adipose - Visceral (Omentum)](javascript:gtex.goTissuePage('Esophagus_Muscularis')) |
|  |  |  |  |  |  |  |  | 7.50E-05 | -0.2 | [Adrenal Gland](javascript:gtex.goTissuePage('Nerve_Tibial')) |
|  |  |  |  |  |  |  |  | 3.80E-12 | -0.2 | [Artery - Aorta](javascript:gtex.goTissuePage('Artery_Tibial')) |
|  |  |  |  |  |  |  |  | 1.30E-06 | -0.19 | [Artery - Coronary](javascript:gtex.goTissuePage('Esophagus_Muscularis')) |
|  |  |  |  |  |  |  |  | 5.00E-27 | -0.22 | [Artery - Tibial](javascript:gtex.goTissuePage('Heart_Atrial_Appendage')) |
|  |  |  |  |  |  |  |  | 1.10E-04 | -0.16 | [Brain - Caudate (basal ganglia)](javascript:gtex.goTissuePage('Thyroid')) |
|  |  |  |  |  |  |  |  | 4.00E-07 | -0.24 | [Brain - Cerebellar Hemisphere](javascript:gtex.goTissuePage('Esophagus_Gastroesophageal_Junction')) |
|  |  |  |  |  |  |  |  | 3.90E-05 | -0.16 | [Brain - Cerebellum](javascript:gtex.goTissuePage('Skin_Not_Sun_Exposed_Suprapubic')) |
|  |  |  |  |  |  |  |  | 1.40E-05 | -0.1 | [Breast - Mammary Tissue](javascript:gtex.goTissuePage('Lung')) |
|  |  |  |  |  |  |  |  | 6.00E-07 | -0.15 | [Cells - Cultured fibroblasts](javascript:gtex.goTissuePage('Colon_Sigmoid')) |
|  |  |  |  |  |  |  |  | 1.80E-06 | -0.44 | [Cells - EBV-transformed lymphocytes](javascript:gtex.goTissuePage('Artery_Aorta')) |
|  |  |  |  |  |  |  |  | 8.30E-09 | -0.19 | [Esophagus - Mucosa](javascript:gtex.goTissuePage('Lung')) |
|  |  |  |  |  |  |  |  | 2.80E-11 | -0.14 | [Esophagus - Muscularis](javascript:gtex.goTissuePage('Whole_Blood')) |
|  |  |  |  |  |  |  |  | 1.90E-12 | -0.22 | [Heart - Atrial Appendage](javascript:gtex.goTissuePage('Thyroid')) |
|  |  |  |  |  |  |  |  | 1.40E-09 | -0.15 | [Lung](javascript:gtex.goTissuePage('Esophagus_Mucosa')) |
|  |  |  |  |  |  |  |  | 3.00E-20 | -0.23 | [Nerve - Tibial](javascript:gtex.goTissuePage('Nerve_Tibial')) |
|  |  |  |  |  |  |  |  | 1.50E-08 | -0.27 | [Pancreas](javascript:gtex.goTissuePage('Adipose_Subcutaneous')) |
|  |  |  |  |  |  |  |  | 8.10E-07 | -0.27 | [Pituitary](javascript:gtex.goTissuePage('Pancreas')) |
|  |  |  |  |  |  |  |  | 7.20E-08 | -0.21 | [Prostate](javascript:gtex.goTissuePage('Skin_Sun_Exposed_Lower_leg')) |
|  |  |  |  |  |  |  |  | 8.70E-05 | -0.11 | [Skin - Not Sun Exposed (Suprapubic)](javascript:gtex.goTissuePage('Esophagus_Mucosa')) |
|  |  |  |  |  |  |  |  | 2.20E-08 | -0.15 | [Skin - Sun Exposed (Lower leg)](javascript:gtex.goTissuePage('Adipose_Visceral_Omentum')) |
|  |  |  |  |  |  |  |  | 8.40E-05 | -0.1 | [Testis](javascript:gtex.goTissuePage('Adrenal_Gland')) |
|  |  |  |  |  |  |  |  | 5.30E-11 | -0.19 | [Thyroid](javascript:gtex.goTissuePage('Skin_Not_Sun_Exposed_Suprapubic')) |
|  |  |  |  |  |  |  | RP11-545A16.1 | 5.60E-05 | -0.16 | [Colon - Transverse](javascript:gtex.goTissuePage('Skin_Sun_Exposed_Lower_leg')) |
|  |  |  |  |  |  |  | RP11-545A16.3 | 1.10E-04 | 0.12 | [Artery - Tibial](javascript:gtex.goTissuePage('Prostate')) |
|  |  |  |  |  |  |  | SOAT1 | 1.40E-07 | -0.25 | [Adrenal Gland](javascript:gtex.goTissuePage('Lung')) |
|  |  |  |  |  |  |  |  | 1.70E-11 | 0.15 | [Artery - Tibial](javascript:gtex.goTissuePage('Brain_Cerebellar_Hemisphere')) |
|  |  |  |  |  |  |  |  | 2.00E-04 | -0.13 | [Testis](javascript:gtex.goTissuePage('Cells_Cultured_fibroblasts')) |
|  |  |  |  |  |  |  | TDRD5 | 2.50E-08 | 0.29 | [Adipose - Subcutaneous](javascript:gtex.goTissuePage('Artery_Aorta')) |
|  |  |  |  |  |  |  |  | 2.20E-06 | 0.3 | [Adipose - Visceral (Omentum)](javascript:gtex.goTissuePage('Pituitary')) |
|  |  |  |  |  |  |  |  | 1.40E-06 | 0.33 | [Artery - Aorta](javascript:gtex.goTissuePage('Brain_Cerebellar_Hemisphere')) |
|  |  |  |  |  |  |  |  | 3.20E-11 | 0.32 | [Artery - Tibial](javascript:gtex.goTissuePage('Brain_Frontal_Cortex_BA9')) |
|  |  |  |  |  |  |  |  | 1.70E-05 | 0.49 | [Brain - Cerebellar Hemisphere](javascript:gtex.goTissuePage('Pituitary')) |
|  |  |  |  |  |  |  |  | 1.40E-05 | 0.37 | [Brain - Cortex](javascript:gtex.goTissuePage('Breast_Mammary_Tissue')) |
|  |  |  |  |  |  |  |  | 4.90E-05 | 0.38 | [Brain - Frontal Cortex (BA9)](javascript:gtex.goTissuePage('Pancreas')) |
|  |  |  |  |  |  |  |  | 1.20E-05 | 0.42 | [Brain - Hypothalamus](javascript:gtex.goTissuePage('Adipose_Visceral_Omentum')) |
|  |  |  |  |  |  |  |  | 5.50E-12 | 0.45 | [Esophagus - Mucosa](javascript:gtex.goTissuePage('Artery_Coronary')) |
|  |  |  |  |  |  |  |  | 2.80E-10 | 0.36 | [Lung](javascript:gtex.goTissuePage('Adipose_Subcutaneous')) |
|  |  |  |  |  |  |  |  | 6.80E-09 | 0.35 | [Nerve - Tibial](javascript:gtex.goTissuePage('Cells_EBV-transformed_lymphocytes')) |
|  |  |  |  |  |  |  |  | 1.10E-06 | 0.42 | [Pituitary](javascript:gtex.goTissuePage('Colon_Transverse')) |
|  |  |  |  |  |  |  |  | 4.20E-10 | 0.33 | [Skin - Not Sun Exposed (Suprapubic)](javascript:gtex.goTissuePage('Cells_Cultured_fibroblasts')) |
|  |  |  |  |  |  |  |  | 3.90E-08 | 0.29 | [Skin - Sun Exposed (Lower leg)](javascript:gtex.goTissuePage('Colon_Transverse')) |
|  |  |  |  |  |  |  |  | 2.10E-09 | 0.32 | [Thyroid](javascript:gtex.goTissuePage('Breast_Mammary_Tissue')) |
|  |  |  |  |  |  |  | TOR3A | 2.20E-13 | 0.28 | [Adipose - Subcutaneous](javascript:gtex.goTissuePage('Brain_Cortex')) |
|  |  |  |  |  |  |  |  | 9.00E-08 | 0.19 | [Adipose - Visceral (Omentum)](javascript:gtex.goTissuePage('Brain_Hypothalamus')) |
|  |  |  |  |  |  |  |  | 7.00E-10 | 0.24 | [Artery - Aorta](javascript:gtex.goTissuePage('Brain_Hippocampus')) |
|  |  |  |  |  |  |  |  | 4.10E-23 | 0.34 | [Artery - Tibial](javascript:gtex.goTissuePage('Brain_Nucleus_accumbens_basal_ganglia')) |
|  |  |  |  |  |  |  |  | 3.40E-07 | 0.22 | [Breast - Mammary Tissue](javascript:gtex.goTissuePage('Brain_Cerebellar_Hemisphere')) |
|  |  |  |  |  |  |  |  | 3.50E-13 | 0.22 | [Cells - Cultured fibroblasts](javascript:gtex.goTissuePage('Brain_Cortex')) |
|  |  |  |  |  |  |  |  | 7.20E-10 | 0.34 | [Colon - Sigmoid](javascript:gtex.goTissuePage('Brain_Cerebellum')) |
|  |  |  |  |  |  |  |  | 8.60E-07 | 0.15 | [Colon - Transverse](javascript:gtex.goTissuePage('Artery_Coronary')) |
|  |  |  |  |  |  |  |  | 1.30E-10 | 0.31 | [Esophagus - Gastroesophageal Junction](javascript:gtex.goTissuePage('Brain_Frontal_Cortex_BA9')) |
|  |  |  |  |  |  |  |  | 3.50E-09 | 0.17 | [Esophagus - Mucosa](javascript:gtex.goTissuePage('Adipose_Visceral_Omentum')) |
|  |  |  |  |  |  |  |  | 2.70E-12 | 0.28 | [Esophagus - Muscularis](javascript:gtex.goTissuePage('Adrenal_Gland')) |
|  |  |  |  |  |  |  |  | 9.50E-12 | 0.33 | [Heart - Atrial Appendage](javascript:gtex.goTissuePage('Brain_Cerebellum')) |
|  |  |  |  |  |  |  |  | 1.20E-05 | 0.17 | [Heart - Left Ventricle](javascript:gtex.goTissuePage('Brain_Putamen_basal_ganglia')) |
|  |  |  |  |  |  |  |  | 2.90E-07 | 0.17 | [Lung](javascript:gtex.goTissuePage('Testis')) |
|  |  |  |  |  |  |  |  | 4.40E-12 | 0.24 | [Nerve - Tibial](javascript:gtex.goTissuePage('Pancreas')) |
|  |  |  |  |  |  |  |  | 4.20E-07 | 0.25 | [Pancreas](javascript:gtex.goTissuePage('Stomach')) |
|  |  |  |  |  |  |  |  | 6.50E-08 | 0.21 | [Skin - Not Sun Exposed (Suprapubic)](javascript:gtex.goTissuePage('Brain_Frontal_Cortex_BA9')) |
|  |  |  |  |  |  |  |  | 2.90E-16 | 0.31 | [Skin - Sun Exposed (Lower leg)](javascript:gtex.goTissuePage('Heart_Left_Ventricle')) |
|  |  |  |  |  |  |  |  | 1.80E-14 | 0.24 | [Thyroid](javascript:gtex.goTissuePage('Brain_Caudate_basal_ganglia')) |
|  |  |  |  |  |  |  |  | 6.50E-10 | 0.2 | [Whole Blood](javascript:gtex.goTissuePage('Skin_Not_Sun_Exposed_Suprapubic')) |
| Depression  &  Related traits | 2p25.3 | rs13029479 | 2 | 655222 | A | G | AC092159.2 | 7.70E-05 | -0.33 | Adipose - Subcutaneous |
|  | 4q13.3 | rs10013800 | 4 | 71731236 | T | C | GRSF1 | 9.40E-07 | 0.37 | Esophagus - Mucosa |
|  | 19q13.32 | rs4802269 | 19 | 46167469 | A | G | DMPK | 9.30E-05 | 0.085 | Adipose - Subcutaneous |
|  |  |  |  |  |  |  |  | 5.70E-07 | 0.13 | Nerve - Tibial |
|  |  |  |  |  |  |  |  | 5.00E-05 | 0.096 | Skin - Sun Exposed (Lower leg) |
|  |  |  |  |  |  |  |  | 2.90E-06 | 0.12 | Thyroid |
|  |  |  |  |  |  |  | DMWD | 2.50E-06 | 0.13 | Adipose - Subcutaneous |
|  |  |  |  |  |  |  |  | 2.50E-05 | 0.16 | Artery - Aorta |
|  |  |  |  |  |  |  |  | 6.20E-07 | 0.13 | Artery - Tibial |
|  |  |  |  |  |  |  | EML2 | 3.60E-05 | -0.25 | Brain - Cerebellar Hemisphere |
|  |  |  |  |  |  |  | EML2-AS1 | 2.30E-06 | -0.18 | Whole Blood |
|  |  |  |  |  |  |  | GIPR | 1.20E-05 | 0.17 | Adipose - Visceral (Omentum) |
|  |  |  |  |  |  |  | QPCTL | 1.50E-05 | 0.11 | Nerve - Tibial |
|  | 2q37.1 | rs12993249 | 2 | 234479377 | A | G | DGKD | 4.70E-05 | 0.1 | Nerve - Tibial |
|  |  |  |  |  |  |  |  | 4.80E-05 | 0.081 | Skin - Sun Exposed (Lower leg) |
|  |  |  |  |  |  |  | USP40 | 1.70E-09 | -0.16 | Adipose - Subcutaneous |
|  |  |  |  |  |  |  |  | 1.10E-07 | -0.13 | Artery - Tibial |
|  |  |  |  |  |  |  |  | 2.30E-06 | -0.2 | Brain - Anterior cingulate cortex (BA24) |
|  |  |  |  |  |  |  |  | 4.90E-05 | -0.26 | Brain - Cerebellar Hemisphere |
|  |  |  |  |  |  |  |  | 4.10E-13 | -0.43 | Brain - Cerebellum |
|  |  |  |  |  |  |  |  | 1.50E-09 | -0.2 | Brain - Cortex |
|  |  |  |  |  |  |  |  | 3.50E-07 | -0.21 | Brain - Frontal Cortex (BA9) |
|  |  |  |  |  |  |  |  | 1.50E-06 | -0.2 | Brain - Hypothalamus |
|  |  |  |  |  |  |  |  | 3.80E-05 | -0.14 | Breast - Mammary Tissue |
|  |  |  |  |  |  |  |  | 2.00E-26 | -0.26 | Cells - Cultured fibroblasts |
|  |  |  |  |  |  |  |  | 2.10E-05 | -0.19 | Colon - Transverse |
|  |  |  |  |  |  |  |  | 7.70E-08 | -0.17 | Esophagus - Mucosa |
|  |  |  |  |  |  |  |  | 4.50E-12 | -0.25 | Heart - Atrial Appendage |
|  |  |  |  |  |  |  |  | 2.50E-06 | -0.18 | Heart - Left Ventricle |
|  |  |  |  |  |  |  |  | 6.70E-07 | -0.28 | Pituitary |
|  |  |  |  |  |  |  |  | 6.90E-05 | -0.1 | Skin - Not Sun Exposed (Suprapubic) |
|  |  |  |  |  |  |  |  | 1.20E-05 | -0.11 | Skin - Sun Exposed (Lower leg) |
|  |  |  |  |  |  |  |  | 2.70E-05 | -0.15 | Stomach |
|  |  |  |  |  |  |  |  | 5.90E-11 | -0.17 | Thyroid |
|  |  |  |  |  |  |  |  | 5.30E-05 | 0.12 | Whole Blood |
|  | 4q13.3 | rs6850072 | 4 | 71725868 | A | G | DCK | 1.00E-06 | 0.25 | Artery - Aorta |
|  |  |  |  |  |  |  |  | 8.10E-06 | 0.2 | Artery - Tibial |
|  |  |  |  |  |  |  |  | 1.20E-05 | 0.12 | Cells - Cultured fibroblasts |
|  |  |  |  |  |  |  |  | 2.90E-09 | 0.27 | Esophagus - Mucosa |
|  |  |  |  |  |  |  |  | 3.20E-05 | 0.18 | Muscle - Skeletal |
|  |  |  |  |  |  |  |  | 1.30E-04 | 0.13 | Skin - Not Sun Exposed (Suprapubic) |
|  |  |  |  |  |  |  |  | 9.00E-05 | 0.15 | Skin - Sun Exposed (Lower leg) |
|  |  |  |  |  |  |  |  | 1.90E-05 | 0.2 | Thyroid |
|  |  |  |  |  |  |  | RUFY3 | 1.60E-06 | -0.21 | Adipose - Subcutaneous |
|  |  |  |  |  |  |  |  | 8.00E-05 | -0.2 | Artery - Aorta |
|  |  |  |  |  |  |  |  | 4.20E-07 | -0.21 | Artery - Tibial |
|  |  |  |  |  |  |  |  | 4.50E-06 | -0.22 | Breast - Mammary Tissue |
|  |  |  |  |  |  |  |  | 1.20E-04 | -0.2 | Colon - Transverse |
|  |  |  |  |  |  |  |  | 1.80E-08 | -0.25 | Esophagus - Mucosa |
|  |  |  |  |  |  |  |  | 1.00E-05 | -0.21 | Esophagus - Muscularis |
|  |  |  |  |  |  |  |  | 3.00E-15 | -0.36 | Muscle - Skeletal |
|  |  |  |  |  |  |  |  | 9.10E-12 | -0.3 | Nerve - Tibial |
|  |  |  |  |  |  |  |  | 1.50E-05 | -0.22 | Testis |
| Schizophrenia  &  Related traits | 5q31.3 | rs11956411 | 5 | 140853856 | C | T | FCHSD1 | 8.70E-06 | -0.12 | Cells - Cultured fibroblasts |
|  |  |  |  |  |  |  | PCDHGA2 | 3.90E-11 | -0.36 | Adipose - Subcutaneous |
|  |  |  |  |  |  |  |  | 4.20E-06 | -0.3 | Adipose - Visceral (Omentum) |
|  |  |  |  |  |  |  |  | 1.60E-05 | -0.21 | Artery - Tibial |
|  |  |  |  |  |  |  |  | 3.80E-05 | -0.29 | Colon - Sigmoid |
|  |  |  |  |  |  |  |  | 2.10E-06 | -0.25 | Esophagus - Muscularis |
|  |  |  |  |  |  |  |  | 1.90E-07 | -0.26 | Nerve - Tibial |
|  |  |  |  |  |  |  |  | 6.30E-05 | -0.27 | Pituitary |
|  |  |  |  |  |  |  |  | 1.20E-04 | -0.21 | Skin - Sun Exposed (Lower leg) |
|  |  |  |  |  |  |  | PCDHGA3 | 9.60E-05 | 0.24 | Brain - Cortex |
|  |  |  |  |  |  |  | PCDHGA8 | 2.40E-04 | -0.22 | Cells - Cultured fibroblasts |
|  |  |  |  |  |  |  | PCDHGB5 | 3.50E-05 | 0.16 | Artery - Tibial |
|  |  |  |  |  |  |  |  | 4.60E-05 | 0.17 | Thyroid |
|  |  |  |  |  |  |  | PCDHGC5 | 8.50E-05 | -0.15 | Artery - Tibial |
|  |  |  |  |  |  |  |  | 8.90E-06 | -0.19 | Colon - Transverse |
|  |  |  |  |  |  |  |  | 1.90E-06 | -0.26 | Heart - Atrial Appendage |
|  |  |  |  |  |  |  | RELL2 | 6.10E-06 | 0.21 | Thyroid |
|  | 5q31.3 | rs2237079 | 5 | 140877905 | G | A | ARAP3 | 1.70E-06 | -0.09 | Whole Blood |
|  |  |  |  |  |  |  | DIAPH1 | 2.90E-05 | -0.09 | Artery - Tibial |
|  |  |  |  |  |  |  |  | 2.70E-05 | -0.09 | Esophagus - Muscularis |
|  |  |  |  |  |  |  | FCHSD1 | 8.00E-06 | 0.091 | Esophagus - Muscularis |
|  |  |  |  |  |  |  | HDAC3 | 1.00E-05 | 0.11 | Esophagus - Mucosa |
|  |  |  |  |  |  |  |  | 3.00E-05 | 0.091 | Skin - Not Sun Exposed (Suprapubic) |
|  |  |  |  |  |  |  | PCDHA10 | 1.40E-04 | -0.17 | Nerve - Tibial |
|  |  |  |  |  |  |  | PCDHA9 | 1.90E-04 | -0.21 | Nerve - Tibial |
|  |  |  |  |  |  |  | PCDHGA10 | 1.60E-04 | 0.097 | Cells - Cultured fibroblasts |
|  |  |  |  |  |  |  | PCDHGA8 | 2.00E-05 | -0.2 | Cells - Cultured fibroblasts |
|  | 4q13.1 | rs62297721 | 4 | 66504803 | A | G | EPHA5 | 6.40E-09 | 0.25 | Nerve - Tibial |
|  |  |  |  |  |  |  |  | 2.60E-20 | 0.47 | Thyroid |
|  |  |  |  |  |  |  | EPHA5-AS1 | 8.50E-13 | 0.41 | Thyroid |
|  |  |  |  |  |  |  | RP11-807H7.2 | 9.30E-12 | 0.38 | Thyroid |
|  | 8p21.2 | rs117325001 | 8 | 26242272 | G | T | BNIP3L | 4.60E-09 | -0.14 | Esophagus - Mucosa |
|  |  |  |  |  |  |  |  | 4.90E-05 | -0.13 | Nerve - Tibial |
|  |  |  |  |  |  |  | SDAD1P1 | 1.00E-29 | -0.66 | Adipose - Subcutaneous |
|  |  |  |  |  |  |  |  | 1.20E-16 | -0.45 | Adipose - Visceral (Omentum) |
|  |  |  |  |  |  |  |  | 1.00E-13 | -0.68 | Adrenal Gland |
|  |  |  |  |  |  |  |  | 4.40E-23 | -0.58 | Artery - Aorta |
|  |  |  |  |  |  |  |  | 1.30E-13 | -0.64 | Artery - Coronary |
|  |  |  |  |  |  |  |  | 1.50E-29 | -0.65 | Artery - Tibial |
|  |  |  |  |  |  |  |  | 1.00E-06 | -0.5 | Brain - Caudate (basal ganglia) |
|  |  |  |  |  |  |  |  | 1.30E-06 | -0.67 | Brain - Cerebellar Hemisphere |
|  |  |  |  |  |  |  |  | 9.00E-07 | -0.63 | Brain - Cerebellum |
|  |  |  |  |  |  |  |  | 6.00E-07 | -0.6 | Brain - Putamen (basal ganglia) |
|  |  |  |  |  |  |  |  | 7.60E-16 | -0.55 | Breast - Mammary Tissue |
|  |  |  |  |  |  |  |  | 6.90E-32 | -0.53 | Cells - Cultured fibroblasts |
|  |  |  |  |  |  |  |  | 4.30E-14 | -0.69 | Cells - EBV-transformed lymphocytes |
|  |  |  |  |  |  |  |  | 4.00E-16 | -0.6 | Colon - Sigmoid |
|  |  |  |  |  |  |  |  | 1.40E-18 | -0.42 | Colon - Transverse |
|  |  |  |  |  |  |  |  | 5.00E-20 | -0.6 | Esophagus - Gastroesophageal Junction |
|  |  |  |  |  |  |  |  | 2.20E-17 | -0.47 | Esophagus - Mucosa |
|  |  |  |  |  |  |  |  | 7.50E-33 | -0.64 | Esophagus - Muscularis |
|  |  |  |  |  |  |  |  | 3.00E-22 | -0.52 | Heart - Atrial Appendage |
|  |  |  |  |  |  |  |  | 5.10E-16 | -0.41 | Heart - Left Ventricle |
|  |  |  |  |  |  |  |  | 2.20E-19 | -0.41 | Lung |
|  |  |  |  |  |  |  |  | 2.00E-05 | -0.48 | Minor Salivary Gland |
|  |  |  |  |  |  |  |  | 7.40E-54 | -0.72 | Muscle - Skeletal |
|  |  |  |  |  |  |  |  | 3.20E-23 | -0.54 | Nerve - Tibial |
|  |  |  |  |  |  |  |  | 1.20E-10 | -0.59 | Ovary |
|  |  |  |  |  |  |  |  | 3.20E-16 | -0.64 | Pancreas |
|  |  |  |  |  |  |  |  | 3.80E-09 | -0.47 | Pituitary |
|  |  |  |  |  |  |  |  | 5.10E-11 | -0.56 | Prostate |
|  |  |  |  |  |  |  |  | 2.00E-16 | -0.57 | Skin - Not Sun Exposed (Suprapubic) |
|  |  |  |  |  |  |  |  | 1.80E-29 | -0.69 | Skin - Sun Exposed (Lower leg) |
|  |  |  |  |  |  |  |  | 7.00E-08 | -0.37 | Small Intestine - Terminal Ileum |
|  |  |  |  |  |  |  |  | 8.10E-09 | -0.52 | Spleen |
|  |  |  |  |  |  |  |  | 9.00E-18 | -0.51 | Stomach |
|  |  |  |  |  |  |  |  | 7.70E-15 | -0.38 | Testis |
|  |  |  |  |  |  |  |  | 1.20E-27 | -0.54 | Thyroid |
|  |  |  |  |  |  |  |  | 2.40E-08 | -0.56 | Uterus |
|  |  |  |  |  |  |  |  | 8.60E-06 | -0.52 | Vagina |
|  |  |  |  |  |  |  |  | 1.70E-06 | -0.27 | Whole Blood |
|  | 1q25.2 | rs61824368 | 1 | 179298809 | C | T | ABL2 | 5.50E-05 | 0.33 | Brain - Anterior cingulate cortex (BA24) |
|  |  |  |  |  |  |  |  | 8.30E-07 | 0.31 | Brain - Cerebellar Hemisphere |
|  |  |  |  |  |  |  |  | 2.50E-05 | 0.27 | Brain - Cerebellum |
|  |  |  |  |  |  |  |  | 1.10E-05 | 0.32 | Brain - Cortex |
|  |  |  |  |  |  |  |  | 7.10E-07 | 0.34 | Brain - Frontal Cortex (BA9) |
|  |  |  |  |  |  |  |  | 1.40E-05 | 0.28 | Brain - Hippocampus |
|  |  |  |  |  |  |  |  | 1.60E-05 | 0.18 | Brain - Nucleus accumbens (basal ganglia) |
|  |  |  |  |  |  |  |  | 3.10E-05 | 0.23 | Brain - Putamen (basal ganglia) |
|  |  |  |  |  |  |  |  | 3.00E-06 | 0.12 | Cells - Cultured fibroblasts |
|  |  |  |  |  |  |  | AXDND1 | 1.90E-06 | -0.2 | Colon - Transverse |
|  |  |  |  |  |  |  | FAM20B | 9.30E-07 | -0.11 | Adipose - Subcutaneous |
|  |  |  |  |  |  |  |  | 4.40E-05 | -0.13 | Adipose - Visceral (Omentum) |
|  |  |  |  |  |  |  |  | 7.50E-05 | -0.2 | Adrenal Gland |
|  |  |  |  |  |  |  |  | 3.80E-12 | -0.2 | Artery - Aorta |
|  |  |  |  |  |  |  |  | 1.30E-06 | -0.19 | Artery - Coronary |
|  |  |  |  |  |  |  |  | 5.00E-27 | -0.22 | Artery - Tibial |
|  |  |  |  |  |  |  |  | 1.10E-04 | -0.16 | Brain - Caudate (basal ganglia) |
|  |  |  |  |  |  |  |  | 4.00E-07 | -0.24 | Brain - Cerebellar Hemisphere |
|  |  |  |  |  |  |  |  | 3.90E-05 | -0.16 | Brain - Cerebellum |
|  |  |  |  |  |  |  |  | 1.40E-05 | -0.1 | Breast - Mammary Tissue |
|  |  |  |  |  |  |  |  | 6.00E-07 | -0.15 | Cells - Cultured fibroblasts |
|  |  |  |  |  |  |  |  | 1.80E-06 | -0.44 | Cells - EBV-transformed lymphocytes |
|  |  |  |  |  |  |  |  | 8.30E-09 | -0.19 | Esophagus - Mucosa |
|  |  |  |  |  |  |  |  | 2.80E-11 | -0.14 | Esophagus - Muscularis |
|  |  |  |  |  |  |  |  | 1.90E-12 | -0.22 | Heart - Atrial Appendage |
|  |  |  |  |  |  |  |  | 1.40E-09 | -0.15 | Lung |
|  |  |  |  |  |  |  |  | 3.00E-20 | -0.23 | Nerve - Tibial |
|  |  |  |  |  |  |  |  | 1.50E-08 | -0.27 | Pancreas |
|  |  |  |  |  |  |  |  | 8.10E-07 | -0.27 | Pituitary |
|  |  |  |  |  |  |  |  | 7.20E-08 | -0.21 | Prostate |
|  |  |  |  |  |  |  |  | 8.70E-05 | -0.11 | Skin - Not Sun Exposed (Suprapubic) |
|  |  |  |  |  |  |  |  | 2.20E-08 | -0.15 | Skin - Sun Exposed (Lower leg) |
|  |  |  |  |  |  |  |  | 8.40E-05 | -0.1 | Testis |
|  |  |  |  |  |  |  |  | 5.30E-11 | -0.19 | Thyroid |
|  |  |  |  |  |  |  | RP11-545A16.1 | 5.60E-05 | -0.16 | Colon - Transverse |
|  |  |  |  |  |  |  | RP11-545A16.3 | 1.10E-04 | 0.12 | Artery - Tibial |
|  |  |  |  |  |  |  | SOAT1 | 1.40E-07 | -0.25 | Adrenal Gland |
|  |  |  |  |  |  |  |  | 1.70E-11 | 0.15 | Artery - Tibial |
|  |  |  |  |  |  |  |  | 2.00E-04 | -0.13 | Testis |
|  |  |  |  |  |  |  | TDRD5 | 2.50E-08 | 0.29 | Adipose - Subcutaneous |
|  |  |  |  |  |  |  |  | 2.20E-06 | 0.3 | Adipose - Visceral (Omentum) |
|  |  |  |  |  |  |  |  | 1.40E-06 | 0.33 | Artery - Aorta |
|  |  |  |  |  |  |  |  | 3.20E-11 | 0.32 | Artery - Tibial |
|  |  |  |  |  |  |  |  | 1.70E-05 | 0.49 | Brain - Cerebellar Hemisphere |
|  |  |  |  |  |  |  |  | 1.40E-05 | 0.37 | Brain - Cortex |
|  |  |  |  |  |  |  |  | 4.90E-05 | 0.38 | Brain - Frontal Cortex (BA9) |
|  |  |  |  |  |  |  |  | 1.20E-05 | 0.42 | Brain - Hypothalamus |
|  |  |  |  |  |  |  |  | 5.50E-12 | 0.45 | Esophagus - Mucosa |
|  |  |  |  |  |  |  |  | 2.80E-10 | 0.36 | Lung |
|  |  |  |  |  |  |  |  | 6.80E-09 | 0.35 | Nerve - Tibial |
|  |  |  |  |  |  |  |  | 1.10E-06 | 0.42 | Pituitary |
|  |  |  |  |  |  |  |  | 4.20E-10 | 0.33 | Skin - Not Sun Exposed (Suprapubic) |
|  |  |  |  |  |  |  |  | 3.90E-08 | 0.29 | Skin - Sun Exposed (Lower leg) |
|  |  |  |  |  |  |  |  | 2.10E-09 | 0.32 | Thyroid |
|  |  |  |  |  |  |  | TOR3A | 2.20E-13 | 0.28 | Adipose - Subcutaneous |
|  |  |  |  |  |  |  |  | 9.00E-08 | 0.19 | Adipose - Visceral (Omentum) |
|  |  |  |  |  |  |  |  | 7.00E-10 | 0.24 | Artery - Aorta |
|  |  |  |  |  |  |  |  | 4.10E-23 | 0.34 | Artery - Tibial |
|  |  |  |  |  |  |  |  | 3.40E-07 | 0.22 | Breast - Mammary Tissue |
|  |  |  |  |  |  |  |  | 3.50E-13 | 0.22 | Cells - Cultured fibroblasts |
|  |  |  |  |  |  |  |  | 7.20E-10 | 0.34 | Colon - Sigmoid |
|  |  |  |  |  |  |  |  | 8.60E-07 | 0.15 | Colon - Transverse |
|  |  |  |  |  |  |  |  | 1.30E-10 | 0.31 | Esophagus - Gastroesophageal Junction |
|  |  |  |  |  |  |  |  | 3.50E-09 | 0.17 | Esophagus - Mucosa |
|  |  |  |  |  |  |  |  | 2.70E-12 | 0.28 | Esophagus - Muscularis |
|  |  |  |  |  |  |  |  | 9.50E-12 | 0.33 | Heart - Atrial Appendage |
|  |  |  |  |  |  |  |  | 1.20E-05 | 0.17 | Heart - Left Ventricle |
|  |  |  |  |  |  |  |  | 2.90E-07 | 0.17 | Lung |
|  |  |  |  |  |  |  |  | 4.40E-12 | 0.24 | Nerve - Tibial |
|  |  |  |  |  |  |  |  | 4.20E-07 | 0.25 | Pancreas |
|  |  |  |  |  |  |  |  | 6.50E-08 | 0.21 | Skin - Not Sun Exposed (Suprapubic) |
|  |  |  |  |  |  |  |  | 2.90E-16 | 0.31 | Skin - Sun Exposed (Lower leg) |
|  |  |  |  |  |  |  |  | 1.80E-14 | 0.24 | Thyroid |
|  |  |  |  |  |  |  |  | 6.50E-10 | 0.2 | Whole Blood |

**Table S3. Multi-trait analysis between depression and schizophrenia in East Asians using MTAG.**

| **SNP** | **CHR** | **BP** | **A1** | **A2** | **Depression** | | **Schizophrenia** | | **MTAG(Depression)** | | **Genes** | **Annotation** |
| --- | --- | --- | --- | --- | --- | --- | --- | --- | --- | --- | --- | --- |
|  |  |  |  |  | **beta** | ***P*-value** | **beta** | ***P*-value** | **beta** | ***P*-value** |  |  |
| rs12031894 | 1 | 179270314 | T | C | -0.054 | 9.82E-04 | -0.075 | 1.24E-07 | -0.014 | 1.44E-08 | SOAT1 | new |
| rs17590956 | 2 | 200720420 | A | G | 0.063 | 0.002 | 0.105 | 6.40E-09 | 0.017 | 5.42E-09 | FTCDNL1 | known |
| rs28735056 | 18 | 77622879 | A | G | -0.044 | 0.004 | -0.093 | 1.30E-10 | -0.014 | 1.17E-09 | KCNG2 | known |
| **SNP** | **CHR** | **BP** | **A1** | **A2** | **Depression** | | **Schizophrenia** | | **MTAG(Schizophrenia)** | | **Genes** | **Annotation** |
|  |  |  |  |  | **beta** | ***P*-value** | **beta** | ***P*-value** | **beta** | ***P*-value** |  |  |
| rs12031894 | 1 | 179270314 | T | C | -0.054 | 9.82E-04 | -0.075 | 1.24E-07 | -0.033 | 2.23E-08 | SOAT1 | new |
| rs17590956 | 2 | 200720420 | A | G | 0.063 | 0.002 | 0.105 | 6.40E-09 | 0.038 | 1.06E-09 | FTCDNL1 | known |
| rs28735056 | 18 | 77622879 | A | G | -0.044 | 0.004 | -0.093 | 1.30E-10 | -0.039 | 1.97E-11 | KCNG2 | known |

**Table S4.** Significant genetic correlations between psychiatric disorders and deep phenotypes.

| **p1** | **p2** | **r_g_** | **se** | **z** | **p** |
| --- | --- | --- | --- | --- | --- |
| Depression | Body mass index | 0.236 | 0.071 | 3.344 | 8.00E-04 |
|  | HDL cholesterol | -0.202 | 0.092 | -2.202 | 0.028 |
|  | Serum alkaline phosphatase levels | 0.228 | 0.099 | 2.304 | 0.021 |
|  | Total bilirubin levels | -0.205 | 0.103 | -1.983 | 0.047 |
|  | Weight | 0.160 | 0.062 | 2.599 | 0.009 |
|  | Type 2 diabetes | 0.242 | 0.081 | 2.987 | 0.003 |
|  | Drugs used in diabetes use measurement | 0.194 | 0.085 | 2.297 | 0.022 |
| Schizophrenia | HDL cholesterol | -0.131 | 0.048 | -2.739 | 0.006 |
|  | Total cholesterol levels | -0.116 | 0.049 | -2.364 | 0.018 |
|  | Angina pectoris | 0.129 | 0.060 | 2.155 | 0.031 |
|  | Chronic hepatitis C infection | -0.251 | 0.094 | -2.679 | 0.007 |
|  | Chronic sinusitis | -0.262 | 0.130 | -2.015 | 0.044 |
|  | Stable angina pectoris | 0.117 | 0.050 | 2.320 | 0.020 |
|  | Agents acting on the renin-angiotensin system use measurement | 0.187 | 0.049 | 3.841 | 1.00E-04 |
|  | Non-steroidal anti-inflammatory and antirheumatic product use measurement | 0.291 | 0.116 | 2.510 | 0.012 |
|  | Antihypertensive use measurement | 0.166 | 0.057 | 2.898 | 0.004 |
|  | Antithrombotic agent use measurement | 0.166 | 0.051 | 3.269 | 0.001 |
|  | Beta blocking agent use measurement | 0.191 | 0.064 | 2.973 | 0.003 |
|  | Calcium channel blocker use measurement | 0.114 | 0.045 | 2.520 | 0.012 |
|  | Peptic ulcer and gastro-oesophageal reflux disease (GORD) drug use measurement | 0.343 | 0.113 | 3.025 | 0.003 |
|  | HMG CoA reductase inhibitor use measurement | 0.119 | 0.045 | 2.677 | 0.007 |
|  | Aspirin use measurement | 0.132 | 0.043 | 3.098 | 0.002 |
|  | Vasodilators used in cardiac diseases use measurement | 0.157 | 0.055 | 2.845 | 0.004 |

**Table S5.** Diseases and traits associated with novel identified loci in MTAG.

| **Trait** | **Locus** | **SNP** | **Gene** | **Trait** | **P** | **beta** | **OR** | **Study** |
| --- | --- | --- | --- | --- | --- | --- | --- | --- |
| Depression   &  Schizophrenia | 1q25.2 | rs12031894 | SOAT1 | NA | NA | NA | NA | NA |
| Depression  &  Related traits | 2p25.3 | rs13029479 | TMEM18 | Trouble falling or staying asleep, or sleeping too much | 0.000787 | 0.0203498 | NA | UKB Neale v2 |
|  |  |  |  | Other mental disorder | 0.0014 | -0.0475 | 0.954 | UKB SAIGE (2018) |
|  | 4q13.3 | rs10013800 | GRSF1 | Sleep disorders | 0.00378 | -0.64372 | 0.52533454 | FINNGEN_R6 |
|  | 12p12.2 | rs10770745 | SLCO1B3-SLCO1B7 | NA | NA | NA | NA | NA |
|  | 9p24.2 | rs10974438 | GLIS3 | Mania, hypomania, bipolar or manic-depression \| mental health problems ever diagnosed by a professional | 0.0037756 | -0.1750208 | 0.83943959 | UKB Neale v2 |
|  | 12q24.12 | rs11066015 | ALDH2 | NA | NA | NA | NA | NA |
|  | 16p12.3 | rs11074451 | GP2 | Trouble falling or staying asleep, or sleeping too much | 0.0003938 | 0.0130906 |  | UKB Neale v2 |
|  | 16q12.2 | rs35626903 | AMFR | Psychological or psychiatric problem | 0.0048 | -0.0015165 | 0.99848463 | PMID:33959723 |
|  | 2q37.1 | rs12993249 | USP40 | Anxiety disorders | 0.000532 | -0.0605 | 0.94129377 | UKB SAIGE |
|  |  |  |  | Anxiety disorder | 0.000669 | -0.062 | 0.93988289 | UKB SAIGE |
|  | 19q13.32 | rs4802269 | GIPR | Alzheimer's disease or family history of Alzheimer's disease | 2.24E-05 | 0.04355309 |  | PMID:33589840 |
|  |  |  |  | Alzheimer's disease (late onset) | 0.0001287 | 0.0766 | 1.07961015 | PMID:24162737 |
|  | 18p11.22 | rs662872 | MTCL1 | NA | NA | NA | NA | NA |
|  | 4q13.3 | rs6850072 | GRSF1 | Dependent personality disorder | 0.000778 | 0.64467 | 1.90535816 | FINNGEN_R6 |
| Schizophrenia  &  Related traits | 5q31.3 | rs11956411 | PCDHGC3 | Schizophrenia vs Tourette's syndrome and other tic disorders (ordinary least squares (OLS)) | 0.000039 | -0.015 | 0.98511194 | PMID:33686288 |
|  |  |  |  | Obsessive compulsive disorder vs Tourette's syndrome and other tic disorders (ordinary least squares (OLS)) | 0.0022 | -0.0071 | 0.99292515 | PMID:33686288 |
|  |  |  |  | Schizophrenia vs major depressive disorder (ordinary least squares (OLS)) | 0.002 | -0.0081 | 0.99193272 | PMID:33686288 |
|  |  |  |  | Schizophrenia vs autism spectrum disorder (ordinary least squares (OLS)) | 0.0046 | -0.0085 | 0.99153602 | PMID:33686288 |
|  | 5q31.3 | rs2237079 | PCDHGC5 | Post-traumatic stress disorder | 0.00301 | 0.12009 | 1.12759833 | FINNGEN_R6 |
|  |  |  |  | Depression | 0.0019 | -0.0014687 | 0.99853241 | PMID:33959723 |
|  |  |  |  | Psychological or psychiatric problem | 0.00028 | -0.0019411 | 0.99806076 | PMID:33959723 |
|  | 4q13.1 | rs62297721 | EPHA5 | Mental and behavioural disorders due to opioids | 0.0012 | 0.19462 | 1.21484926 | FINNGEN_R6 |
|  | 8p21.2 | rs117325001 | BNIP3L | Bipolar disorder vs Tourette's syndrome and other tic disorders (ordinary least squares (OLS)) | 0.0013 | 0.013 | 1.01308487 | PMID:33686288 |
|  |  |  |  | Bipolar disorder vs major depressive disorder (ordinary least squares (OLS)) | 0.0026 | 0.0083 | 1.00833454 | PMID:33686288 |
|  |  |  |  | Schizophrenia vs Tourette's syndrome and other tic disorders (ordinary least squares (OLS)) | 8.8E-07 | 0.018 | 1.01816298 | PMID:33686288 |
|  |  |  |  | Schizophrenia vs obsessive compulsive disorder (ordinary least squares (OLS)) | 0.000047 | 0.013 | 1.01308487 | PMID:33686288 |
|  |  |  |  | Bipolar disorder vs ADHD (ordinary least squares (OLS)) | 0.0017 | 0.011 | 1.01106072 | PMID:33686288 |
|  |  |  |  | Schizophrenia vs autism spectrum disorder (ordinary least squares (OLS)) | 0.000013 | 0.013 | 1.01308487 | PMID:33686288 |
|  |  |  |  | Schizophrenia vs major depressive disorder (ordinary least squares (OLS)) | 4.1E-08 | 0.015 | 1.01511306 | PMID:33686288 |
|  |  |  |  | Schizophrenia vs anorexia nervosa (ordinary least squares (OLS)) | 0.0000055 | 0.015 | 1.01511306 | PMID:33686288 |
|  |  |  |  | Schizophrenia vs ADHD (ordinary least squares (OLS)) | 5.6E-07 | 0.017 | 1.01714532 | PMID:33686288 |
|  | 1q25.2 | rs61824368 | SOAT1 | NA | NA | NA | NA | NA |

**Table S6.** Known loci in multi-trait meta-analysis between schizophrenia and risk factors in East Asians.

| **Trait** | **SNP** | **CHR** | **BP** | **A1** | **A2** | **Trait1** | | **Trait2** | | **MTAG** | | **Genes** | **Annotation** |
| --- | --- | --- | --- | --- | --- | --- | --- | --- | --- | --- | --- | --- | --- |
|  |  |  |  |  |  | **beta** | ***P*-value** | **beta** | ***P*-value** | **beta** | ***P*-value** |  |  |
| HDL | rs4660761 | 1 | 44440146 | A | G | -0.09 | 5.08E-09 | 0.013 | 8.20E-03 | -0.034 | 1.57E-08 | ATP6V0B | known |
|  | rs1518395 | 2 | 58208074 | G | A | 0.112 | 6.71E-14 | -0.014 | 9.80E-03 | 0.044 | 2.70E-13 | VRK2 | known |
|  | rs2282762 | 3 | 50514612 | G | A | 0.088 | 4.11E-10 | 0.014 | 3.70E-03 | 0.038 | 8.42E-11 | CACNA2D2 | known |
|  | rs11191355 | 10 | 104392497 | C | T | -0.104 | 1.65E-11 | 0.013 | 6.60E-03 | -0.038 | 6.99E-11 | TRIM8 | known |
|  | rs1984658 | 12 | 123483426 | A | G | -0.106 | 8.62E-14 | -0.028 | 4.20E-08 | -0.046 | 4.05E-15 | ABCB9 | known |
|  | rs9567393 | 13 | 32763757 | G | A | -0.091 | 1.13E-09 | 0.015 | 6.90E-03 | -0.035 | 3.97E-09 | BRCA2 | known |
| TCL | rs60527016 | 8 | 38299624 | T | C | 0.077 | 1.79E-07 | 0.004 | 7.30E-03 | 0.033 | 3.40E-08 | FGFR1 | known |
|  | rs1984658 | 12 | 123483426 | A | G | -0.106 | 8.62E-14 | 0.004 | 1.50E-06 | -0.047 | 1.93E-15 | ABCB9 | known |
| Angina pectoris | rs2236988 | 3 | 50504896 | A | G | 0.09 | 6.10E-11 | 0.013 | 9.38E-03 | 0.038 | 1.09E-10 | CACNA2D2 | known |
|  | rs4147157 | 10 | 104536360 | A | G | -0.113 | 1.32E-15 | 0.013 | 5.29E-04 | -0.046 | 3.97E-15 | WBP1L | known |
| Chronic hepatitis  C infection | rs77483950 | 3 | 50440490 | A | G | 0.11 | 6.84E-10 | 0.026 | 7.29E-03 | 0.037 | 2.59E-09 | TMEM115 | known |
| Chronic sinusitis | rs146289884 | 6 | 165074400 | T | C | -0.108 | 1.50E-09 | 0.027 | 7.71E-03 | -0.036 | 3.61E-09 | C6orf118 | known |
|  | rs320707 | 7 | 137053273 | A | C | -0.091 | 2.81E-09 | 0.023 | 4.59E-03 | -0.036 | 9.20E-10 | PTN | known |
|  | rs7909591 | 10 | 104659018 | T | G | -0.113 | 3.52E-13 | 0.022 | 6.15E-03 | -0.044 | 7.40E-14 | CNNM2 | known |
| Stable angina  pectoris | rs2236988 | 3 | 50504896 | A | G | 0.09 | 6.10E-11 | 0.012 | 1.18E-03 | 0.038 | 1.42E-10 | CACNA2D2 | known |
|  | rs7909591 | 10 | 104659018 | T | G | -0.113 | 3.52E-13 | 0.012 | 1.05E-04 | -0.042 | 1.08E-12 | CNNM2 | known |
| Agents acting  on the renin-angiotensin  system use measurement | rs78138837 | 3 | 50344178 | A | G | -0.101 | 2.59E-11 | 0.008 | 9.31E-03 | -0.038 | 1.14E-10 | HYAL1 | known |
|  | rs4604479 | 8 | 65312647 | A | G | 0.102 | 1.64E-08 | 0.01 | 2.07E-03 | 0.037 | 3.40E-09 | BHLHE22 | known |
|  | rs7909591 | 10 | 104659018 | T | G | -0.113 | 3.52E-13 | 0.009 | 9.59E-05 | -0.041 | 3.15E-12 | CNNM2 | known |
|  | rs28604484 | 12 | 123856259 | A | G | 0.101 | 3.16E-11 | 0.01 | 7.67E-03 | 0.041 | 7.57E-12 | SBNO1 | known |
| Non-steroidal  anti-inflammatory and antirheumatic product use measurement | rs709210 | 3 | 50357869 | A | C | -0.111 | 1.46E-12 | 0.012 | 2.00E-03 | -0.044 | 2.11E-13 | HYAL2 | known |
|  | rs298182 | 8 | 65303535 | A | G | -0.102 | 1.62E-08 | 0.01 | 1.17E-03 | -0.037 | 2.85E-09 | BHLHE22 | known |
|  | rs4148866 | 12 | 123425575 | T | C | -0.102 | 1.15E-12 | 0.01 | 2.76E-03 | -0.043 | 1.48E-13 | OGFOD2 | known |
| Antihypertensive  use measurement | rs7596038 | 2 | 58383820 | T | C | -0.121 | 1.08E-17 | 0.019 | 2.53E-03 | -0.052 | 9.50E-19 | FANCL | known |
|  | rs62258668 | 3 | 50840549 | A | G | -0.102 | 9.73E-11 | 0.024 | 8.70E-03 | -0.038 | 3.80E-10 | DOCK3 | known |
|  | rs1488935 | 8 | 38133793 | A | G | -0.081 | 3.35E-08 | 0.02 | 5.68E-03 | -0.034 | 9.25E-09 | PLPP5 | known |
|  | rs72841270 | 10 | 104642237 | T | G | 0.106 | 2.88E-12 | 0.022 | 2.61E-03 | 0.041 | 1.58E-11 | AS3MT | known |
|  | rs28735056 | 18 | 77622879 | A | G | -0.093 | 1.30E-10 | 0.02 | 7.37E-03 | -0.039 | 2.64E-11 | KCNG2 | known |
| Antithrombotic agent use measurement | rs2717063 | 2 | 58110969 | A | C | 0.102 | 1.06E-12 | 0.008 | 2.52E-03 | 0.043 | 2.37E-13 | VRK2 | known |
|  | rs62257540 | 3 | 51452763 | A | G | -0.104 | 6.16E-11 | 0.01 | 8.61E-03 | -0.039 | 1.72E-10 | RBM15B | known |
|  | rs72841270 | 10 | 104642237 | T | G | 0.106 | 2.88E-12 | 0.009 | 5.48E-03 | 0.041 | 1.11E-11 | AS3MT | known |
|  | rs897385 | 12 | 123341297 | A | G | 0.084 | 8.45E-09 | 0.01 | 8.90E-03 | 0.033 | 2.21E-08 | HIP1R | known |
| Beta blocking agent use measurement | rs66691851 | 3 | 136154828 | T | C | -0.105 | 9.84E-10 | 0.013 | 7.95E-03 | -0.039 | 1.52E-10 | PCCB | known |
|  | rs4147157 | 10 | 104536360 | A | G | -0.114 | 1.32E-15 | 0.011 | 2.52E-03 | -0.045 | 2.03E-14 | WBP1L | known |
| Calcium channel  blocker use measurement | rs7427564 | 3 | 136274435 | A | G | -0.102 | 3.17E-09 | 0.01 | 7.61E-03 | -0.038 | 9.60E-10 | STAG1 | known |
|  | rs11998709 | 8 | 38252070 | T | C | 0.08 | 3.46E-08 | 0.009 | 5.83E-03 | 0.034 | 1.13E-08 | LETM2 | known |
|  | rs4147157 | 10 | 104536360 | A | G | -0.113 | 1.32E-15 | 0.008 | 3.68E-03 | -0.046 | 6.28E-15 | WBP1L | known |
|  | rs28604484 | 12 | 123856259 | A | G | 0.101 | 3.16E-11 | 0.01 | 5.51E-03 | 0.041 | 8.49E-12 | SBNO1 | known |
|  | rs9908552 | 17 | 1297991 | T | C | 0.075 | 6.76E-08 | 0.008 | 1.01E-03 | 0.033 | 1.58E-08 | YWHAE | known |
| Peptic ulcer and gastro-oesophageal  reflux disease  (GORD) drug use measurement | rs11998709 | 8 | 38252070 | T | C | 0.08 | 3.46E-08 | 0.008 | 1.40E-03 | 0.035 | 7.48E-09 | LETM2 | known |
|  | rs28604484 | 12 | 123856259 | A | G | 0.101 | 3.16E-11 | 0.009 | 8.42E-03 | 0.041 | 7.35E-12 | SBNO1 | known |
| HMG CoA  reductase inhibitor use measurement | rs182317984 | 8 | 65313054 | T | C | -0.102 | 1.63E-08 | 0.011 | 1.15E-03 | -0.036 | 5.32E-09 | BHLHE22 | known |
|  | rs1926032 | 10 | 104829469 | T | C | -0.116 | 7.66E-11 | 0.011 | 3.77E-03 | -0.043 | 1.93E-10 | CNNM2 | known |
| aspirin use  measurement | rs9877046 | 3 | 50352458 | T | G | -0.11 | 2.11E-11 | 0.011 | 8.73E-03 | -0.04 | 5.39E-11 | HYAL1 | known |
|  | rs1926032 | 10 | 104829469 | T | C | -0.116 | 7.66E-11 | 0.01 | 6.98E-03 | -0.043 | 1.92E-10 | CNNM2 | known |
|  | rs3896870 | 12 | 123333611 | T | C | 0.086 | 4.02E-09 | 0.01 | 8.99E-03 | 0.034 | 9.99E-09 | HIP1R | known |
|  | rs10861879 | 12 | 108609634 | A | G | 0.081 | 1.18E-08 | 0.008 | 4.38E-03 | 0.033 | 2.50E-08 | WSCD2 | known |
| Vasodilators used  in cardiac diseases  use measurement | rs2204019 | 2 | 58074053 | A | G | 0.089 | 5.76E-10 | 0.013 | 4.37E-03 | 0.038 | 1.14E-10 | VRK2 | known |
|  | rs4147157 | 10 | 104536360 | A | G | -0.114 | 1.32E-15 | 0.012 | 1.28E-03 | -0.045 | 9.51E-15 | WBP1L | known |

**Figure S1. Manhattan plot of the GWAS meta-analysis between depression and SCZ.**

The green circle shows the results of meta-analysis between depression and SCZ; the orange circle shows the results of multi-trait meta-analysis specific to depression; the purple circle shows the results of multi-trait meta-analysis specific to SCZ. Novel pleiotropic loci are colored in yellow.

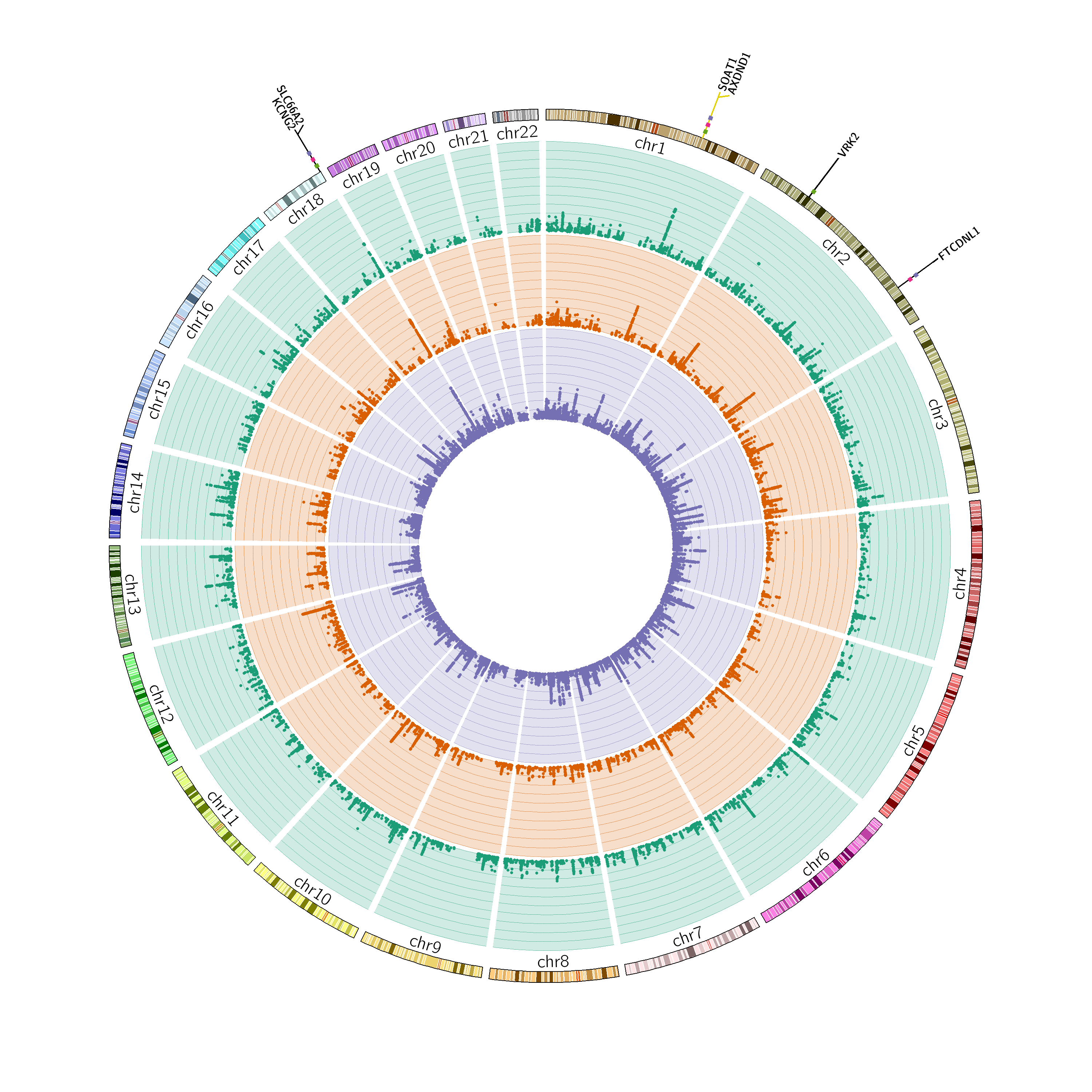

**Figure S2.** Multi-tissue eQTL plot for rs12031894. Multi-tissue eQTL plot for rs12031894. Left panel showed the single-tissue eQTLs for rs12031894, and right panel showed the single-tissue eQTL p-value versus multi-tissue posterior probability. (A) gene *ABL2*; (B) gene *FAM20B*; (C) gene *TDRD5*.

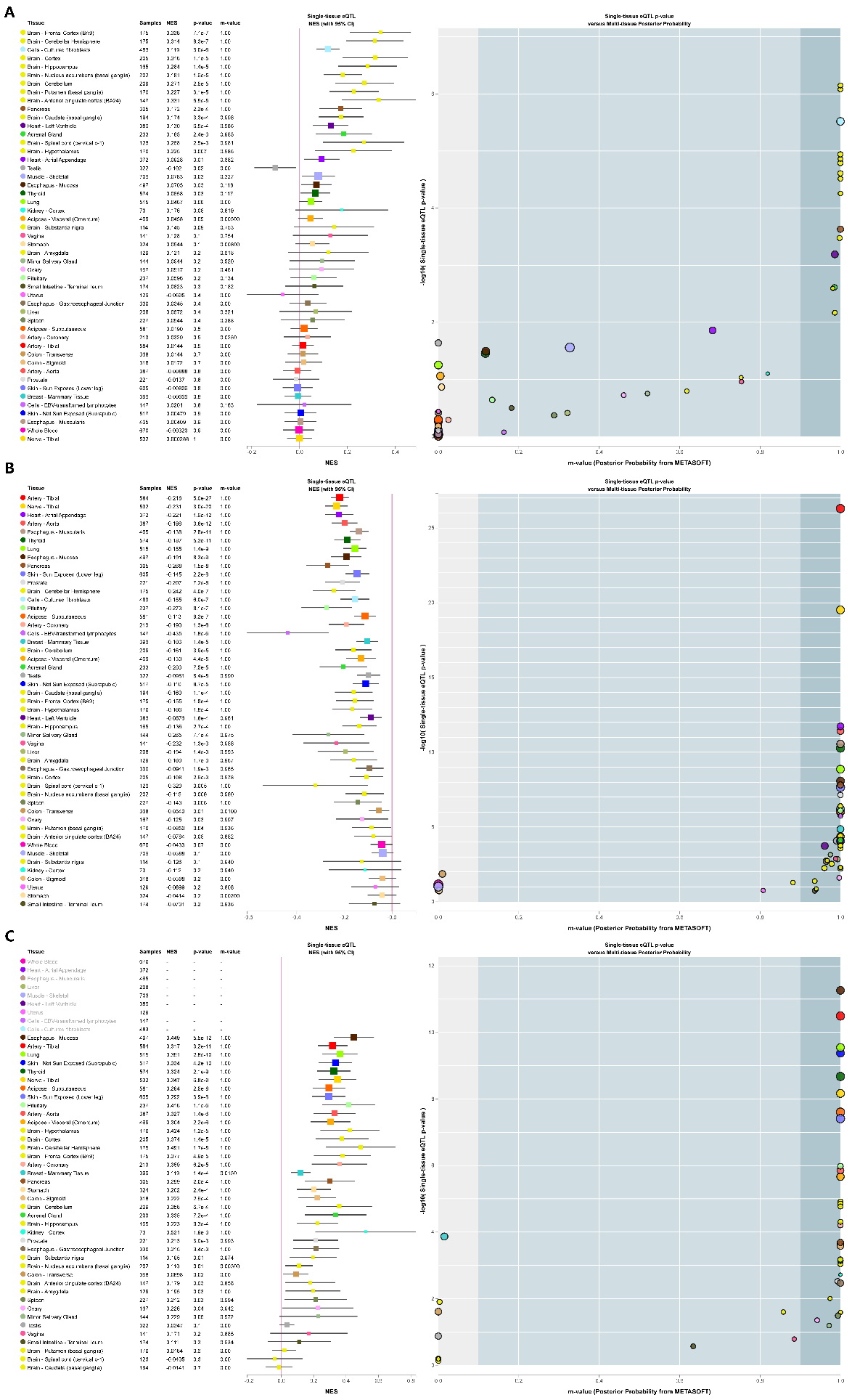

**Figure S3.** Local genetic correlation between psychiatric disorders and HDL cholesterol in East Asians. (A) Manhattan plot showing the estimates of local genetic correlation, genetic covariance, and SNP heritability between depression and HDL cholesterol in East Asians. (B) Manhattan plot showing the estimates of local genetic correlation, genetic covariance, and SNP heritability between schizophrenia and HDL cholesterol in East Asians. Red bars represent loci showing suggestive local genetic correlation (P<0.01). DEP, depression; SCZ, schizophrenia.

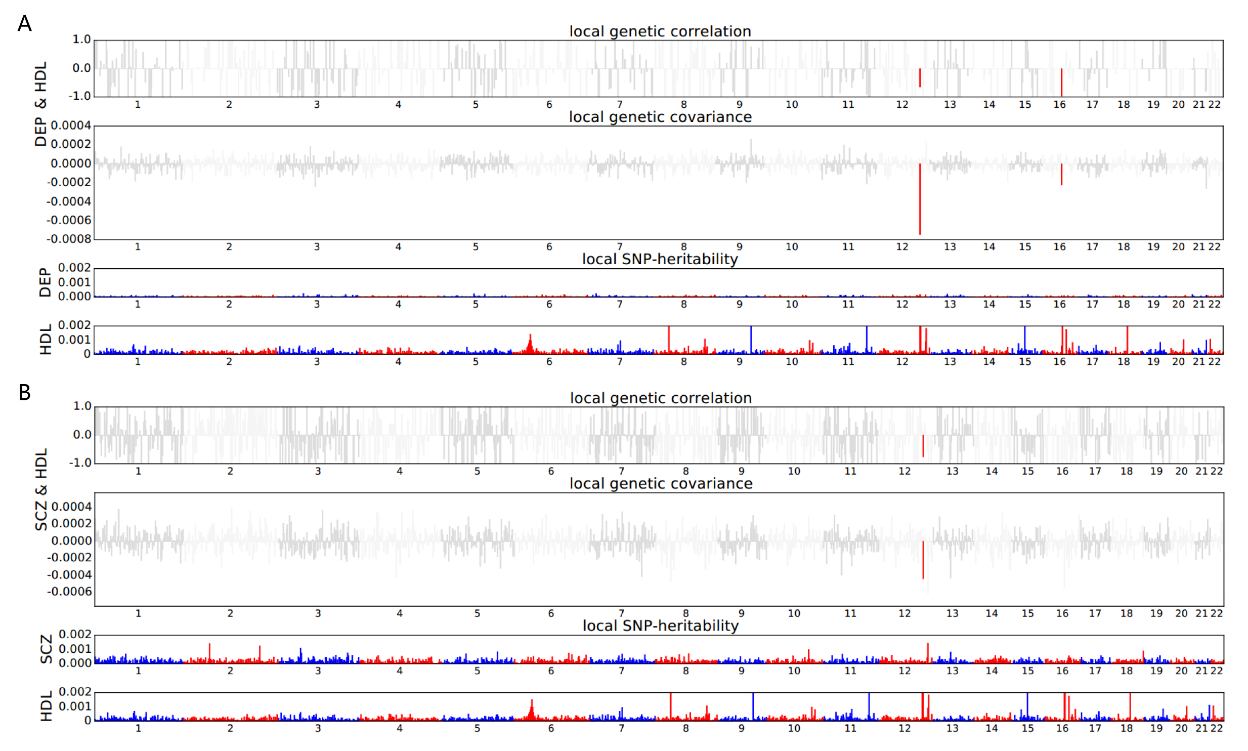

**Figure S4.** Manhattan plot of the GWAS multi-trait meta-analysis between depression and risk factors. The green circle shows the results of multi-trait meta-analysis between depression and BMI; the orange circle shows the results of multi-trait meta-analysis between depression and weight; the purple circle shows the results of multi-trait meta-analysis between depression and HDL cholesterol; the pink circle shows the results of multi-trait meta-analysis between depression and total bilirubin levels; the pink circle shows the results of multi-trait meta-analysis between depression and total bilirubin levels; the light green circle shows the results of multi-trait meta-analysis between depression and type 2 diabetes; the yellow circle shows the results of multi-trait meta-analysis between depression and drugs used in diabetes use measurement. Novel pleiotropic loci are colored in yellow.

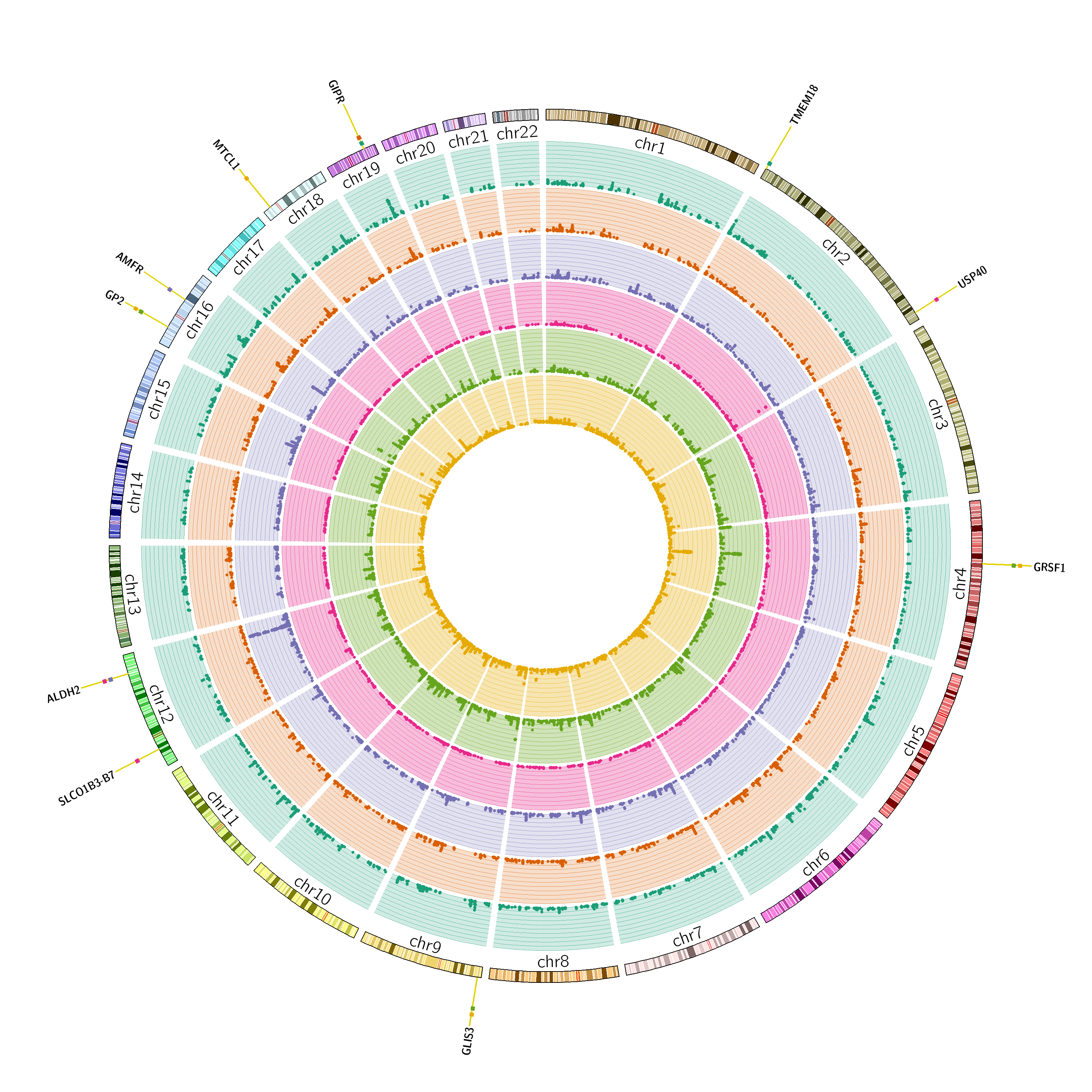

**Figure S5.** Manhattan plot of the GWAS multi-trait meta-analysis between SCZ and risk factors. The green circle shows the results of multi-trait meta-analysis between SCZ and HDL cholesterol; the orange circle shows the results of multi-trait meta-analysis between SCZ and total cholesterol levels; the purple circle shows the results of multi-trait meta-analysis between SCZ and stable angina pectoris; the pink circle shows the results of multi-trait meta-analysis between SCZ and antihypertensive use measurement; the light green circle shows the results of multi-trait meta-analysis between SCZ and antithrombotic agent use measurement; the yellow circle shows the results of multi-trait meta-analysis between SCZ and aspirin use measurement; the khaki circle shows the results of multi-trait meta-analysis between SCZ and vasodilators used in cardiac diseases use measurement. Novel pleiotropic loci are colored in yellow.

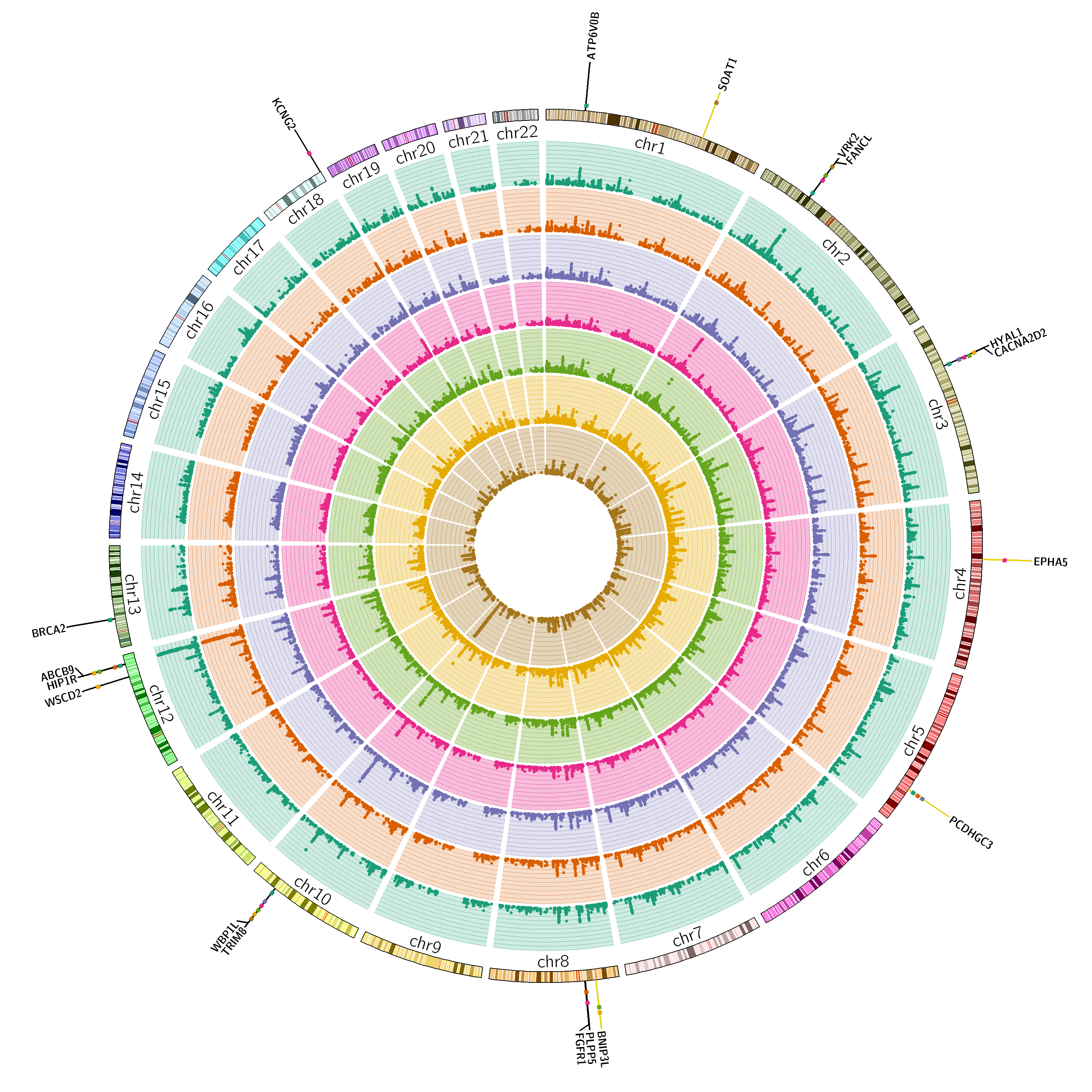

**Figure S6.** Manhattan plot of multi-trait meta-analysis between schizophrenia and angina pectoris in East Asians. Newly identified Loci are depicted in yellow, and previously reported Loci are depicted in blue.

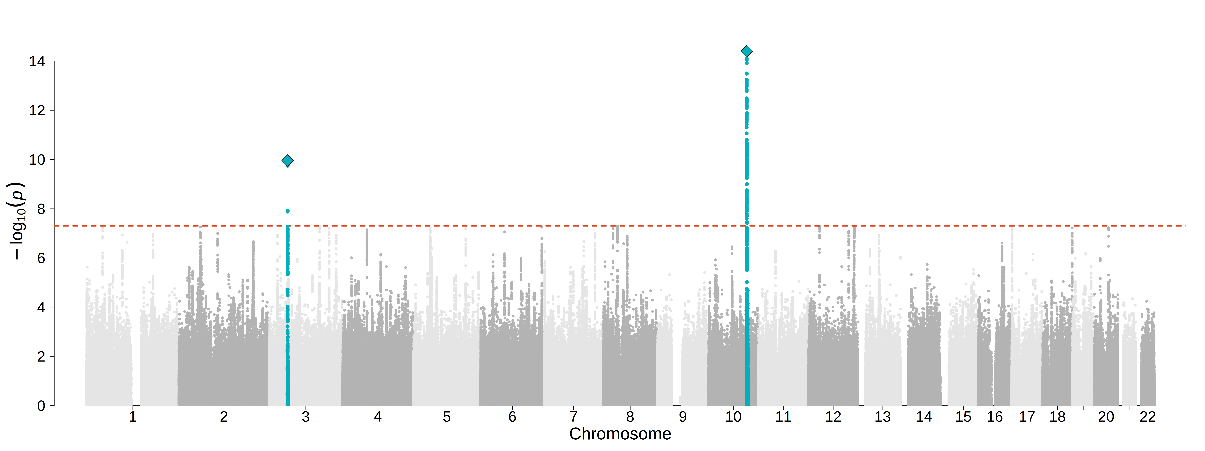

**Figure S7.** Manhattan plot of multi-trait meta-analysis between schizophrenia and chronic hepatitis C infection in East Asians. Newly identified Loci are depicted in yellow, and previously reported Loci are depicted in blue.

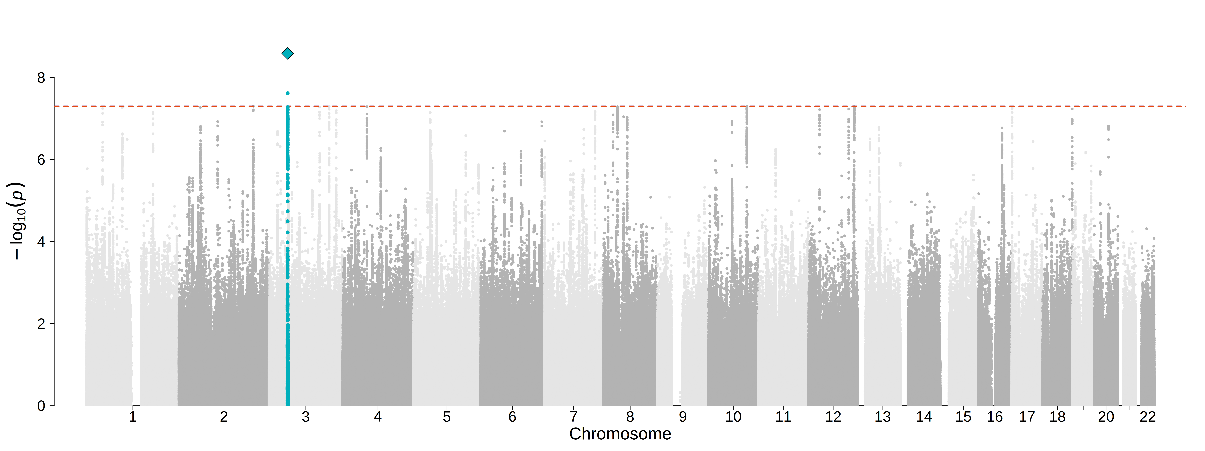

**Figure S8.** Manhattan plot of multi-trait meta-analysis between schizophrenia and chronic sinusitis in East Asians. Newly identified Loci are depicted in yellow, and previously reported Loci are depicted in blue.
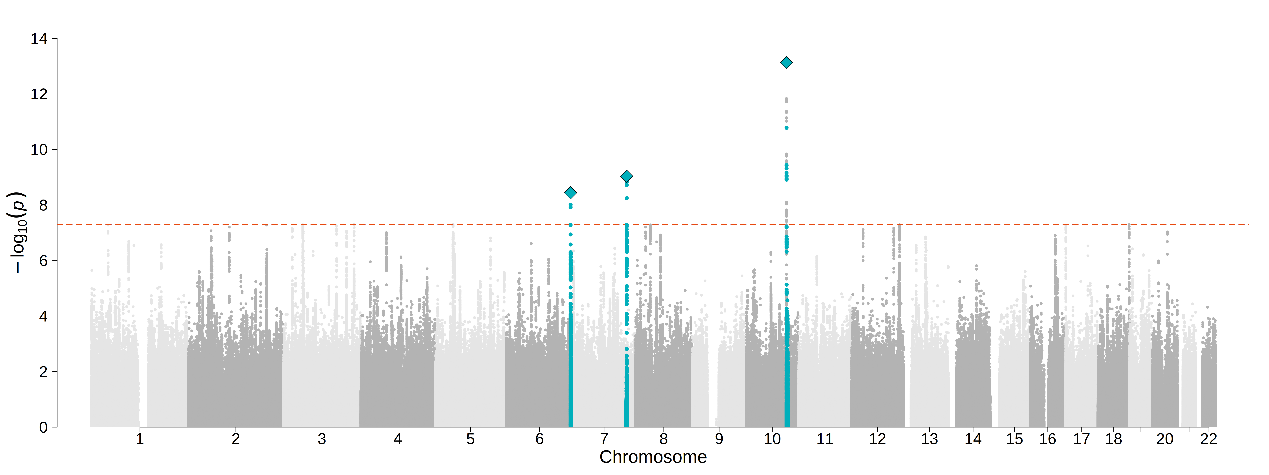

**Figure S9.** Manhattan plot of multi-trait meta-analysis between schizophrenia and agents acting on the renin-angiotensin system use measurement in East Asians. Newly identified Loci are depicted in yellow, and previously reported Loci are depicted in blue.

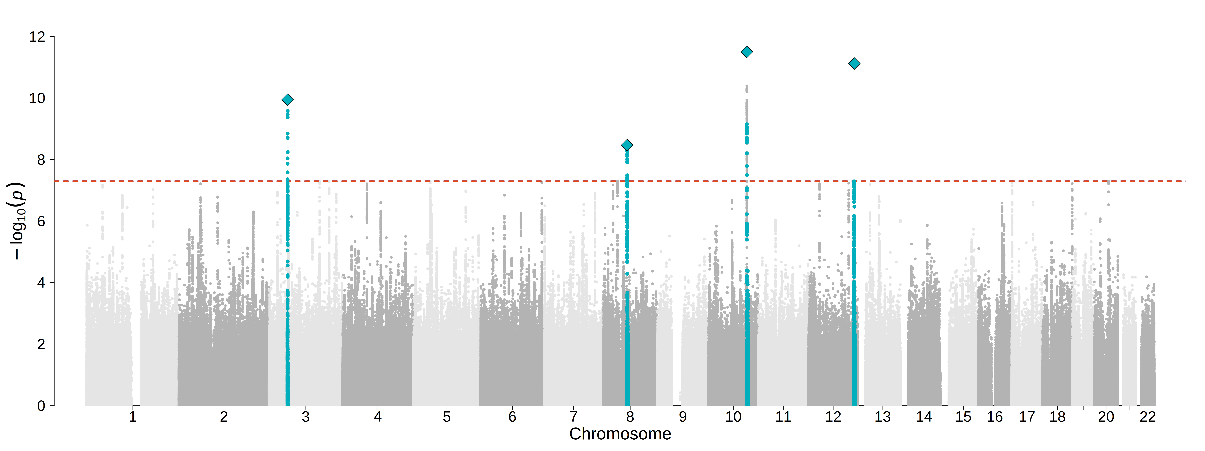

**Figure S10.** Manhattan plot of multi-trait meta-analysis between schizophrenia and non-steroidal anti-inflammatory and antirheumatic product use measurement in East Asians. Newly identified Loci are depicted in yellow, and previously reported Loci are depicted in blue.

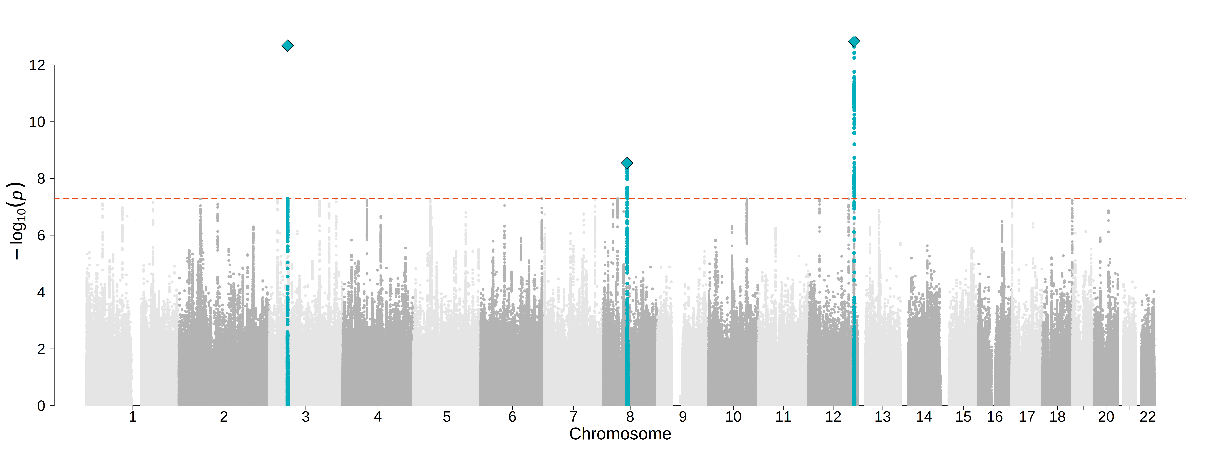

**Figure S11.** Manhattan plot of multi-trait meta-analysis between schizophrenia and beta blocking agent use measurement in East Asians. Newly identified Loci are depicted in yellow, and previously reported Loci are depicted in blue.

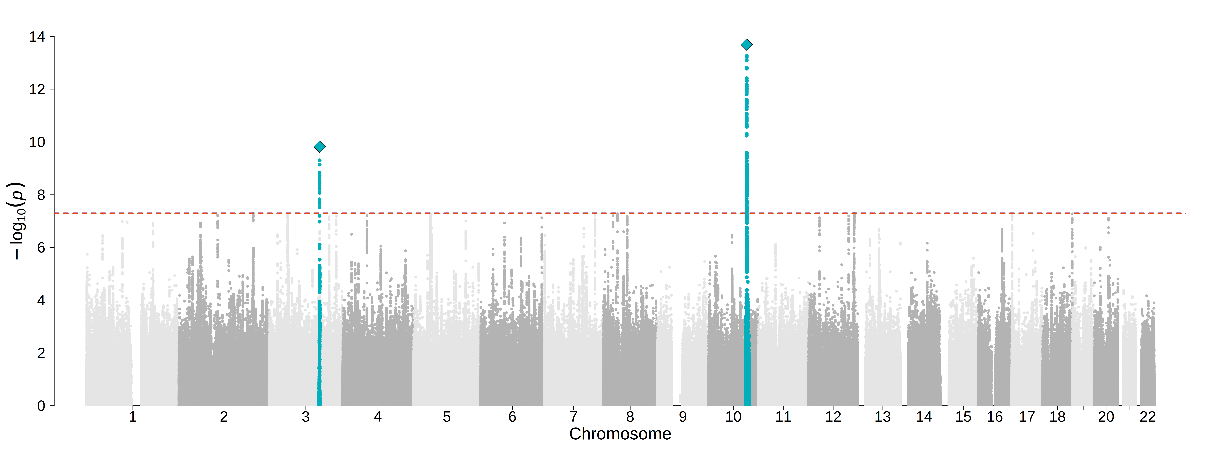

**Figure S12.** Manhattan plot of multi-trait meta-analysis between schizophrenia and calcium channel blocker use measurement in East Asians. Newly identified Loci are depicted in yellow, and previously reported Loci are depicted in blue.

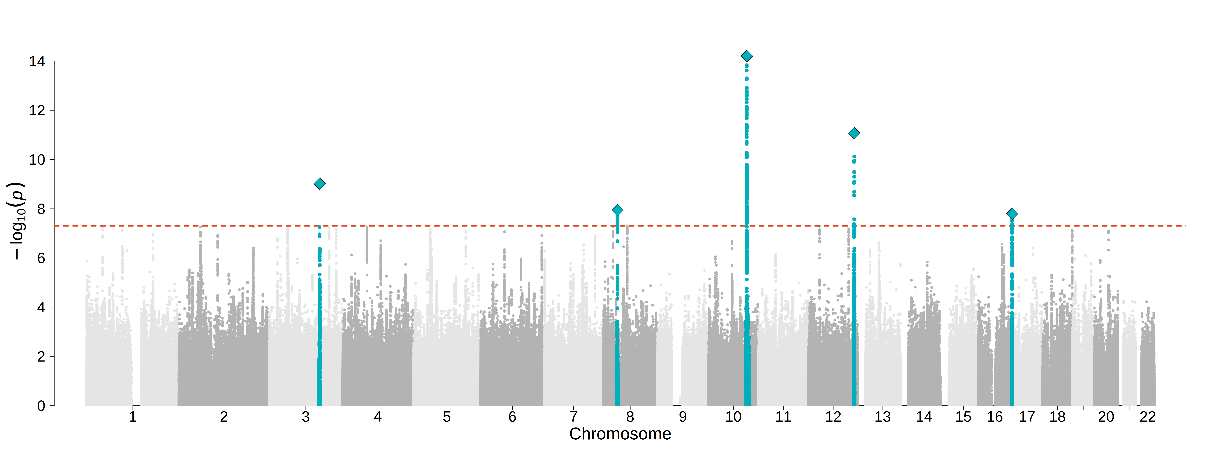

**Figure S13.** Manhattan plot of multi-trait meta-analysis between schizophrenia and peptic ulcer and gastro-oesophageal reflux disease (GORD) drug use measurement in East Asians. Newly identified Loci are depicted in yellow, and previously reported Loci are depicted in blue.

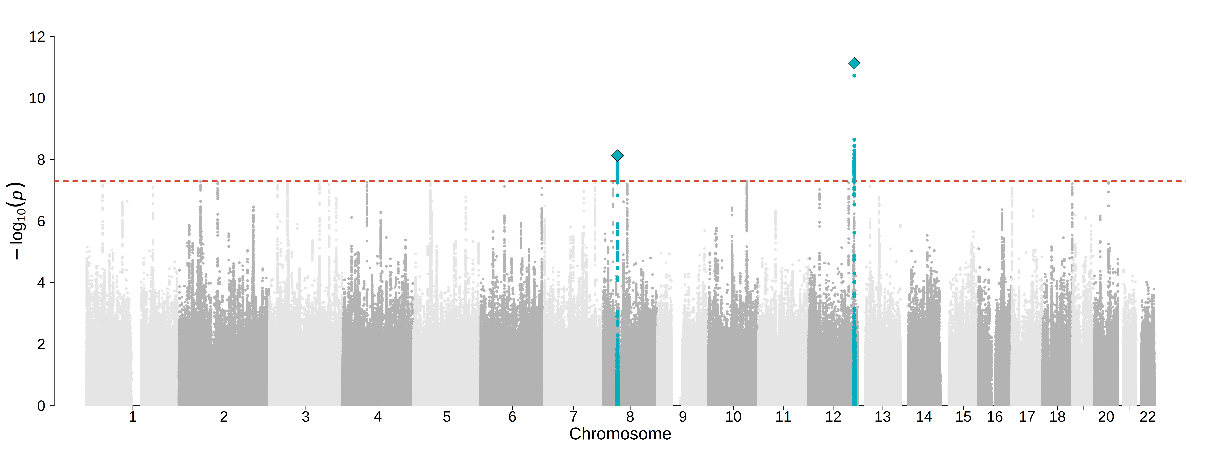

**Figure S14.** Manhattan plot of multi-trait meta-analysis between schizophrenia and HMG CoA reductase inhibitor use measurement in East Asians. Newly identified Loci are depicted in yellow, and previously reported Loci are depicted in blue.

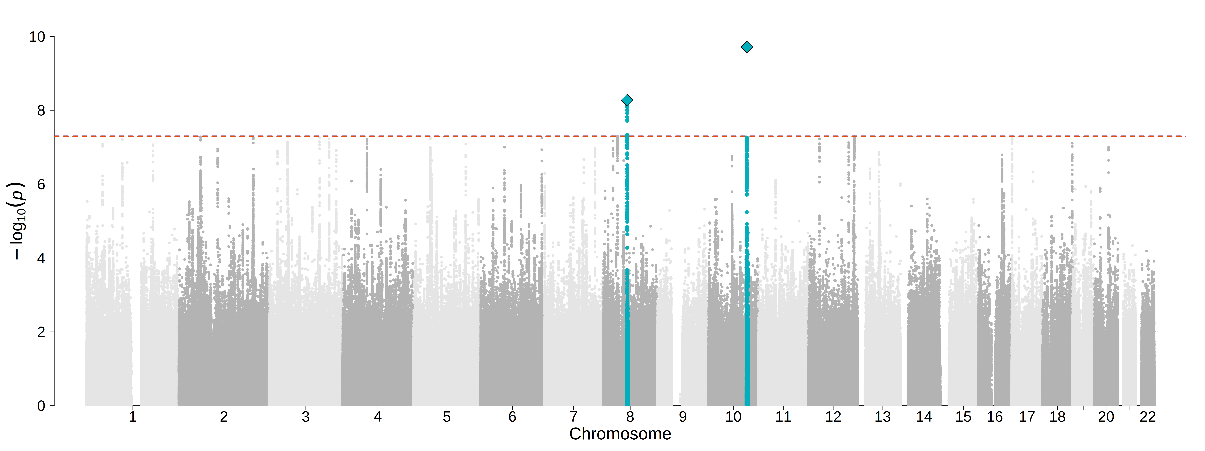
